# Macrophage-targeted glucocorticoid prodrug resolves acute inflammation while preserving HPA axis function: mechanistic, preclinical, and Phase II/III clinical evidence

**DOI:** 10.64898/2026.06.05.26354839

**Authors:** Michael M. Goldberg, Orly Weinreb, Alec M. Goldberg, James I. Goldberg

## Abstract

Glucocorticoids (GCs) remain the fastest-acting anti-inflammatory agents but are constrained by systemic exposure that suppresses the hypothalamic–pituitary–adrenal (HPA) axis, silences adaptive immunity, and drives chronic toxicities. Chronic inflammatory diseases are sustained by long-lived CD206⁺ macrophages containing immune-resistant pathogenic material not cleared physiologically. We developed 101-PGC-005 (’005), a macrophage-targeted type 1a dexamethasone prodrug engineered for low-affinity, recycling-compatible uptake via CD206, with intracellular release triggered by acidic endosomes.

We evaluated ’005 in mechanistic assays, pathogen-diverse preclinical models, three human pharmacokinetic (PK) studies, and an adaptive-design randomized Phase II/III trial in 309 hospitalized patients with moderate COVID-19. In two completed Phase I human studies — a first-in-human dose-escalation and repeated-dose study and a dedicated single-/multiple-dose PK and safety study — ’005 circulated as intact prodrug with rapid systemic clearance (Tmax ∼0.5 h; terminal half-life ∼1.9 h), with no measurable free dexamethasone after single dosing and only low, clinically non-significant free dexamethasone after repeated dosing, and intact prodrug recovered unchanged in urine. Morning cortisol and ACTH were preserved after 30 mg once daily for three consecutive days (1.5× the intended therapeutic dose). A cerebrospinal fluid PK study is evaluating central-compartment penetration.

In the Phase II/III trial — powered for non-inferiority, conducted across six sites in India under GCP with Ministry of Health approval and independent DSMB oversight — ’005 (20 mg IV daily × 3 days) was superior to dexamethasone (6 mg IV daily × 3–10 days) on the primary endpoint of time to ≥2-point improvement on the WHO ordinal scale (HR 2.31; 95% CI 1.83–2.93; p < 0.0001; median 3 vs. 4 days). ’005 was also superior on viral clearance (HR 1.47; 95% CI 1.17–1.84; p = 0.0001), hospital discharge rate, SpO₂ recovery, and fever resolution. Zero patients in the ’005 arm received investigator-initiated corticosteroid supplementation despite protocol allowance. All 309 randomized patients completed the study (ITT = per-protocol). Safety profiles were equivalent (TEAEs 54.8% vs 54.5%; p = 0.958), with no Grade 3+ events, SAEs, deaths, or discontinuations in either arm.

Mechanistically, ’005 delivered dual benefit: acute debulking of inflammatory macrophages and selective depletion of chronically activated pathology-sustaining macrophages, while preserving CXCL10 antiviral signaling and physiologic HPA control. Critically, HPA preservation is not merely a safety feature — it is a core efficacy mechanism: by clearing the pathogenic macrophage burden that was overriding HPA regulation, ’005 restores the conditions for endogenous cortisol to resume its pulsatile, demand-responsive anti-inflammatory role across all GR-expressing cells — lymphocytes, endothelial cells, neurons, and newly differentiated macrophages — that ’005 itself cannot reach. These findings support regulatory-grade evidence for macrophage-targeted corticosteroid therapy and provide the foundation for further development across acute inflammatory indications (sepsis, viral pneumonia, cytokine-release syndromes) and chronic macrophage-driven diseases (atherosclerosis, metabolic steatohepatitis, neurodegeneration, tumor-associated macrophages).

**One Sentence Summary:** A macrophage-targeted dexamethasone prodrug achieves superiority over standard-of-care dexamethasone across two replicate randomized controlled trials totaling 309 patients, without suppressing the hypothalamic–pituitary–adrenal axis.

## Introduction

Inflammation is a whole-body experience, coordinated across immune, neuroendocrine, metabolic, and cardiovascular systems [1]. The HPA axis serves as the central integrator of this response, translating immune stress into finely tuned cortisol release [2]. Through pulsatile circadian and ultradian secretion, cortisol orchestrates systemic adaptation: it engages GC receptors (GRs), expressed in virtually all cells, and mineralocorticoid receptors (MRs), abundant in kidney, heart, and vasculature [3,4].

Under physiologic conditions, cortisol is secreted in circadian and ultradian rhythms that align with sleep–wake cycles, nutrient flux, and immune surveillance. These pulses coordinate not only immune restraint but also vascular tone, glucose handling, neurocognitive performance, and repair processes [5,6]. In acute stress, cortisol release increases rapidly and transiently, mobilizing systemic resources while still maintaining rhythmicity [5]. When inflammation is prolonged, however, cytokines and danger-associated molecular patterns (DAMPs) drive cortisol release outside hypothalamic control, creating mistimed, non-pulsatile exposure [7]. This maladaptive cortisol not only fails to resolve inflammation but downregulates GRs, producing GC resistance [8]. Simultaneously, diversion of cortisol into MR pathways activates the renin–angiotensin–aldosterone system (RAAS), fueling hypertension, insulin resistance, fibrosis, and cardiovascular disease [6,9].

Macrophages sit at the center of this process. These cells are highly elastic: they initiate inflammation by adopting proinflammatory phenotypes and producing cytokines, and they resolve inflammation by transitioning into reparative, tissue-restoring states [10]. Cortisol normally governs this switch, signaling macrophages when to shut down inflammatory programs and enter healing modes.

When HPA control is disrupted, this handoff fails — macrophages remain locked in pathogenic states, and the result is unchecked cytokine storm, tissue injury, and progression to fibrosis [10,11]. In addition, as the last line of defense, macrophages quarantine pathogens that they cannot degrade and other substances such as glucocerebrosidase in Gaucher disease, cholesterol and fat in obesity, metabolic steatohepatitis, atherosclerosis, and peripheral arterial disease, as well as myelin degradation products, amyloid, and tau in central nervous system inflammation [12,13,14,15]. These cells are long-lived, chronically dysregulated, and pro-inflammatory, and are not readily cleared by scavenger macrophages. The ability to drive these cells to apoptosis opens a new pathway to clear immune-resistant material and to treat chronic diseases at their source.

CD206, the mannose receptor, provides a strategic entry point to regulate macrophages. It is expressed on both M1 and M2 macrophages and is dynamically upregulated during inflammation [16]. However, CD206 also exists in soluble form in plasma, creating challenges for therapeutic targeting: high-affinity ligands risk sequestration by this soluble receptor pool, reducing bioavailability and efficacy [17]. Overcoming these challenges requires a design that can exploit CD206 recycling without being trapped in circulation.

Synthetic GCs, introduced in the 1940s, have been indispensable for controlling inflammation, but they fail to replicate the full spectrum of cortisol activities, lack circadian/ultradian rhythm, and suppress endogenous cortisol production [18,19]. Some, such as dexamethasone, do not bind MR, thereby destabilizing RAAS regulation [20,21]. No synthetic GC readily crosses the blood–brain barrier, leaving neuroendocrine regulation unaddressed [22]. Systemic administration overrides the HPA axis, blocking well-controlled cortisol release and producing wide-ranging acute and chronic toxicities [23].

To address these limitations, we developed 101-PGC-005 (’005), a macrophage-targeted dexamethasone prodrug designed to combine the speed and breadth of GC efficacy with preservation of endocrine physiology. Here we report the complete evidence base supporting this approach: mechanistic assays establishing CD206-selective uptake and dual macrophage action; pathogen-diverse preclinical models; two completed Phase I human PK studies confirming prodrug integrity and HPA preservation in vivo, complemented by a cerebrospinal fluid PK study; and a GCP-compliant adaptive Phase II/III randomized controlled trial in 309 patients that achieved superiority over dexamethasone on primary and four secondary endpoints, with zero dropouts and an equivalent safety profile, in two replicate populations separated by over one year across different COVID-19 variant and vaccination landscapes.

## Results

### Design and selective uptake of ’005

’005 was designed as a type 1a dexamethasone prodrug, conjugated through a pH-sensitive oxime-ether linker to a mannose moiety to enable CD206-mediated uptake (Fig. 1A). Unlike high-affinity ligands sequestered by soluble CD206, the low-affinity, recycling-compatible design of ’005 allows repeated receptor engagement and efficient cellular internalization (Fig. 1B). Binding affinity was confirmed using recombinant human CD206 by microscale thermophoresis and surface plasmon resonance (KD = 200.8 µM). Uptake assays with FITC-labeled ’005 demonstrated selective accumulation in primary human macrophages compared with monocytes or lymphocytes, with preferential uptake in CD206⁺ polarized macrophages, with >70% internalization efficiency at 60 minutes (Fig. 1C).

**Fig. 1.**
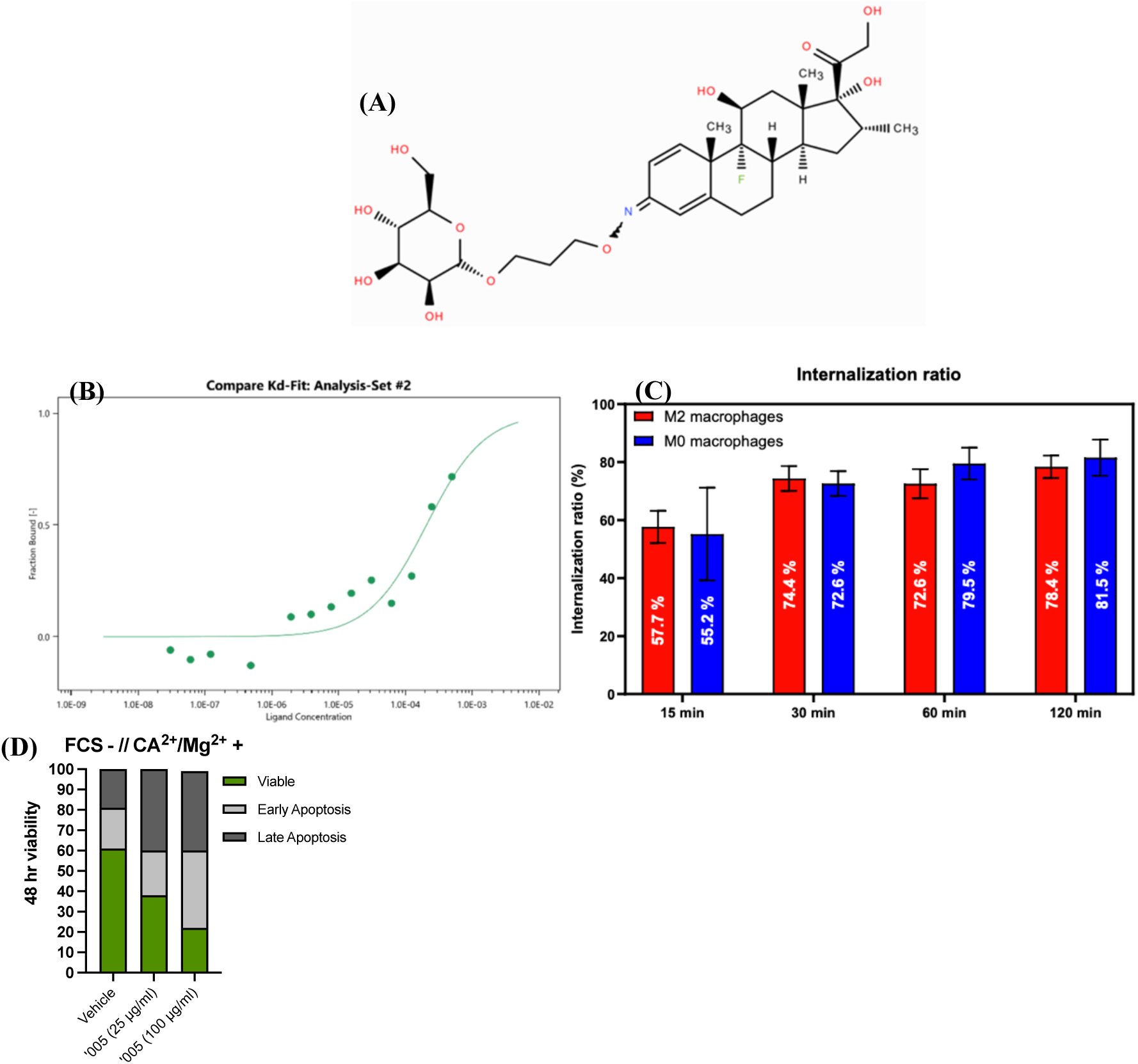
Structure, Binding, Uptake, and Mechanism of Action of ’005. (A) ’005 is comprised of mannose, an oxime-ether linker, and dexamethasone (MW 627 Da). (B) ’005 demonstrated specific binding to recombinant human CD206 with an estimated dissociation constant KD = 200.8 µM. Data shown as fraction bound versus ligand concentration. (C) Trypan blue exclusion assay showing percent internalized FITC-labeled ’005 in human M2 (red) and M0 (blue) macrophages over 120 minutes. Both populations showed >70% internalization efficiency by 60 minutes. (D) Annexin V/PI staining of M2 macrophages treated for 48 hours with ’005 (25 or 100 µg/mL) under serum-free, calcium/magnesium-depleted conditions. Dose-dependent apoptosis observed; equimolar dexamethasone did not induce apoptosis.

Internalization was confirmed by trypan blue quenching and flow cytometry. These results establish that ’005 engages CD206 with appropriate affinity and kinetics to achieve macrophage-specific intracellular delivery.

### Induction of apoptosis in human activated macrophages

To determine whether selective uptake translated into depletion of pathogenic macrophages, we exposed human M2 macrophages to increasing doses of ’005.Peripheral blood mononuclear cells (PBMCs) were isolated from a healthy donor and CD14+ monocytes were enriched using EasySep™ enrichment (StemCell Technologies). Annexin V/propidium iodide staining demonstrated dose-dependent apoptosis at concentrations of 25–100 µg/mL after 48 hours (Fig. 1D). By contrast, dexamethasone at equimolar concentrations did not induce apoptosis, confirming that macrophage depletion is a property unique to targeted intracellular delivery. Vehicle-treated controls showed minimal apoptosis. These results indicate that ’005 reprograms macrophages toward anti-inflammatory states and, at higher intracellular concentrations, induces apoptosis — a dual mechanism of action.

### Cytokine modulation and immune-sparing selectivity

In SARS-CoV-2–infected human macrophages, ’005 rapidly suppressed IL-6 and TNF secretion to levels comparable to dexamethasone but uniquely preserved CXCL10 induction (Fig. 2A–C). CXCL10 is an interferon-inducible chemokine critical for antiviral host defense, suggesting that ’005 spares protective immune signaling while suppressing inflammatory cytokines. This selective profile was consistent across donors (n = 2; IL-6 suppression: two-way ANOVA, p < 0.0001). These results show that ’005 achieves signal selectivity — lowering inflammatory tone while retaining antiviral competence — a property that distinguishes it from both systemic GCs and broad immunosuppressants.

**Fig. 2.**
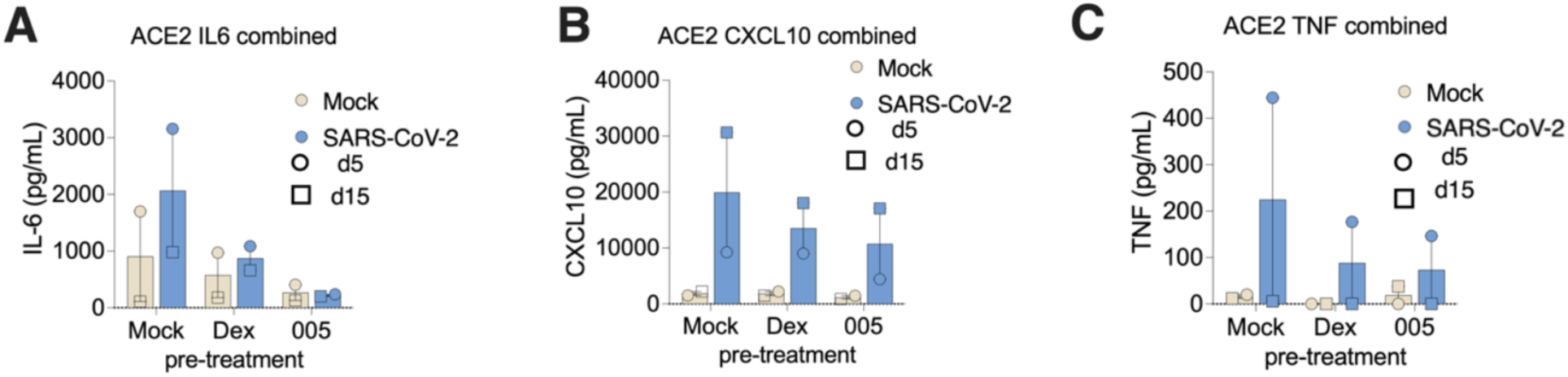
Cytokine modulation in SARS-CoV-2–infected macrophages. (A) IL-6 suppression by ’005 and dexamethasone vs. mock controls in ACE2-expressing human macrophages from two donors (two-way ANOVA, p < 0.0001). (B) CXCL10 preserved by both agents — antiviral signaling not suppressed. (C) TNF suppression in Donor 5; not detectable in Donor 15 under any condition.

### Pathogen-agnostic efficacy across in vivo models

We next evaluated ’005 in pathogen-diverse in vivo models. In an acute LPS challenge in mice, ’005 significantly improved blinded clinical scores at 24 hours compared with dexamethasone or vehicle (Fig. S1). In BALB/c mice infected with Leishmania donovani, ’005 treatment reduced parasite burden in liver and spleen by approximately 8-fold relative to controls (Fig. 3A). In a cutaneous L. major mouse model, repeated dosing prevented ulcer formation and improved lesion scores through day 60 (Fig. 3B). In a bacterial sepsis model (Streptococcus pneumoniae), ’005 lowered peritoneal and blood bacterial loads by 461,000 CFU and 27,000 CFU, respectively (Fig. S2). In a dengue virus mouse model using antibody-dependent enhancement, survival was prolonged in ’005-treated mice compared to controls (Fig. S3) [24]. Together, these findings demonstrate pathogen-agnostic efficacy across bacterial, viral, and parasitic systems, consistent with macrophage-targeted activity rather than pathogen-specific mechanisms.

**Fig. 3.**
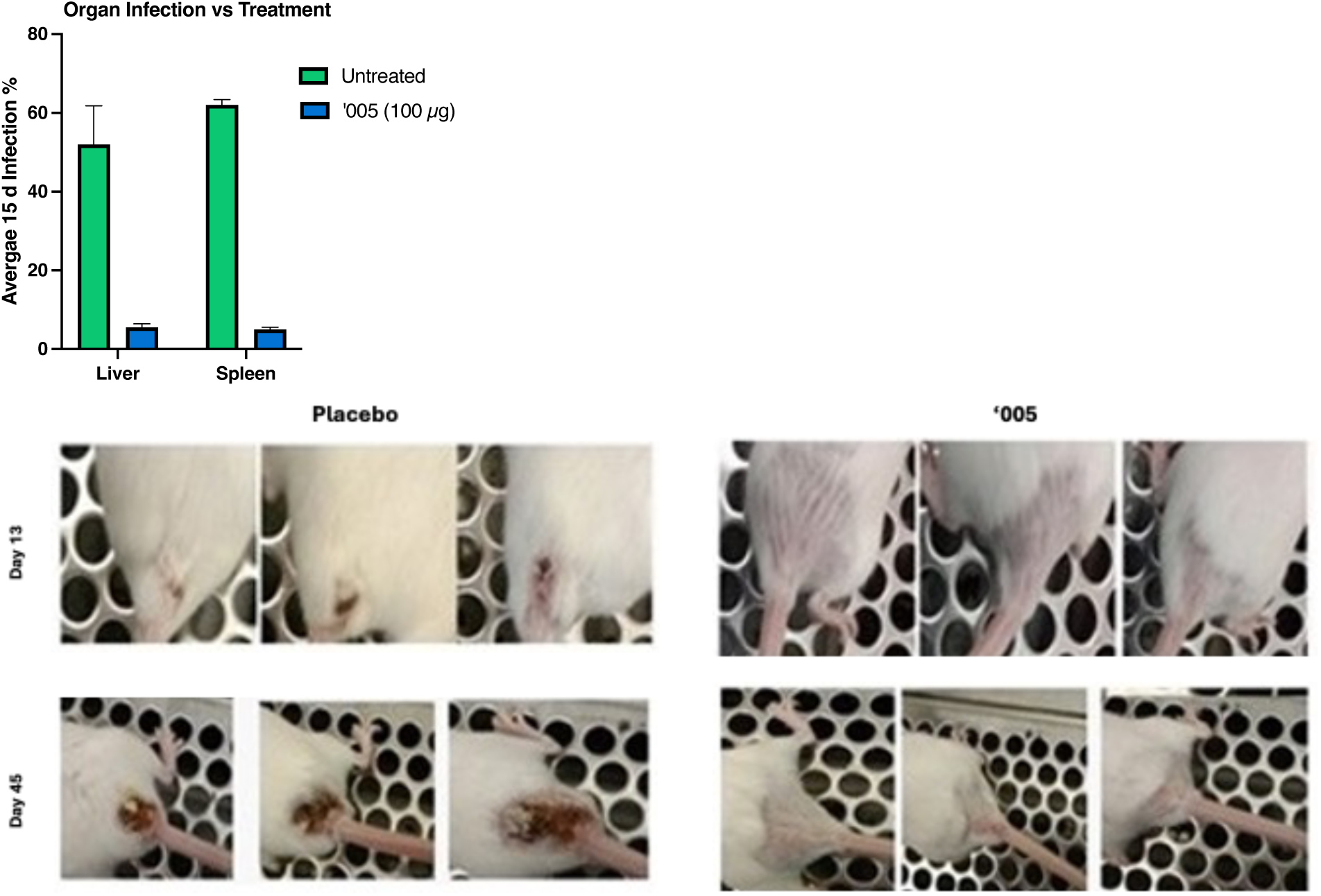
Pathogen-agnostic in vivo efficacy. (A) Organ infection rates in SARS-CoV-2–challenged mice: liver reduced from 57.1% to 7.1%, spleen from 64.3% to 7.1% with ’005 (n=14/group). (B) Anogenital lesion severity in cutaneous leishmaniasis model: severe ulceration in placebo-treated mice absent in ’005-treated animals at Day 13 and Day 45.

### Preclinical toxicology and tissue protection

A 28-day repeat-dose GLP toxicology study in Wistar rats revealed striking differences between ’005 and dexamethasone. Dexamethasone-treated animals exhibited hepatic vacuolation, bone density loss, and pulmonary foamy macrophages, consistent with known GC toxicities. In contrast, 005-treated rats showed dose-dependent and organ-specific histopathologic profiles (Fig. 4A–C). Hepatic vacuolation and pulmonary foamy macrophages were absent or minimal across all ’005 doses, in striking contrast to dexamethasone. Bone density findings were similarly absent or minimal at 10 and 20 mg/kg. At 30 mg/kg, approximately 105× the human mg/kg dose, administered daily for 28 days versus the 3-dose clinical course, 4 of 6 animals exhibited Marked bone density reduction, interpreted as an exaggerated pharmacological effect at extreme supratherapeutic exposure; this finding is discussed in detail below. No bone density changes were observed at 20 mg/kg (∼70× the human mg/kg dose) or below. The NOAEL was 30 mg/kg. Clinical chemistry values remained within normal ranges in all ’005 groups. These results indicate that compartmental restriction of dexamethasone via the prodrug strategy substantially reduces organ toxicity in vivo.

**Fig. 4.**
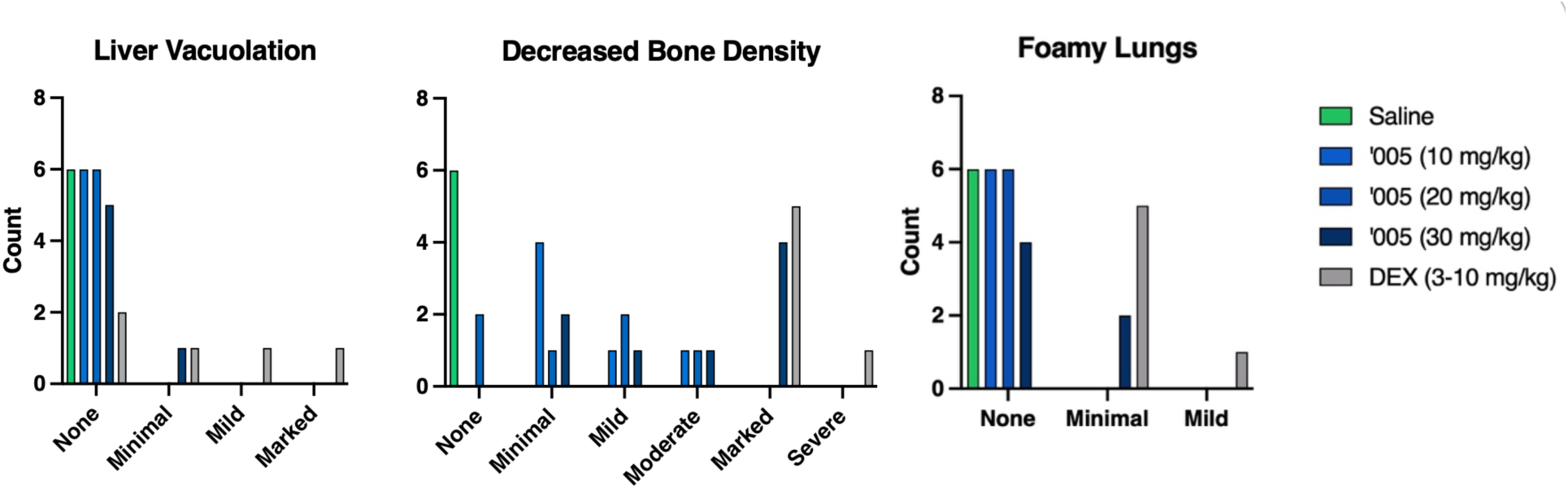
Preclinical toxicology: tissue protection with ’005. 28-day repeat-dose rat study (n = 6/group). (A) Liver vacuolation: mild/minimal in dexamethasone; absent in all ’005 groups. (B) Bone density loss: present in dexamethasone; none to mild in ’005. (C) Foamy lung changes: several dexamethasone animals affected; absent/minimal in ’005.

### Human Pharmacokinetics: Prodrug Integrity and HPA Preservation

A defining question for any type 1a prodrug is whether the active moiety, here, dexamethasone, is released systemically before reaching its intended cellular target. We addressed this in two completed Phase I human studies, a first-in-human dose-escalation and repeated-dose study in Israel and a dedicated single-/multiple-dose PK and safety study in India, complemented by a cerebrospinal fluid (CSF) PK study evaluating central-compartment penetration.

#### Study 1 — First-in-human dose-escalation and repeated-dose study (PIF-GC-CP-101; NCT05619770; Rambam Health Care Campus, Haifa, Israel)

This Phase I, single-center, open-label study enrolled 15 healthy adult volunteers (aged 20–34 years; BMI ≤ 30; both sexes) across a single-dose escalation arm (10, 20, and 30 mg IV bolus) and a repeated-dose arm (30 mg IV once daily for 3 days; n = 3). Free dexamethasone in plasma was very low across all cohorts, with the highest individual value of 0.96 ng/mL, consistent with a stable prodrug that releases only insignificant free dexamethasone systemically. Critically, in the repeated-dose arm, morning cortisol and ACTH measured 24 h after the third dose remained within normal ranges in all three subjects, demonstrating preservation of hypothalamic–pituitary–adrenal (HPA) axis function after three consecutive days of supratherapeutic dosing (Fig. 5). No dose-limiting toxicities or serious adverse events were observed.

**Fig. 5.**
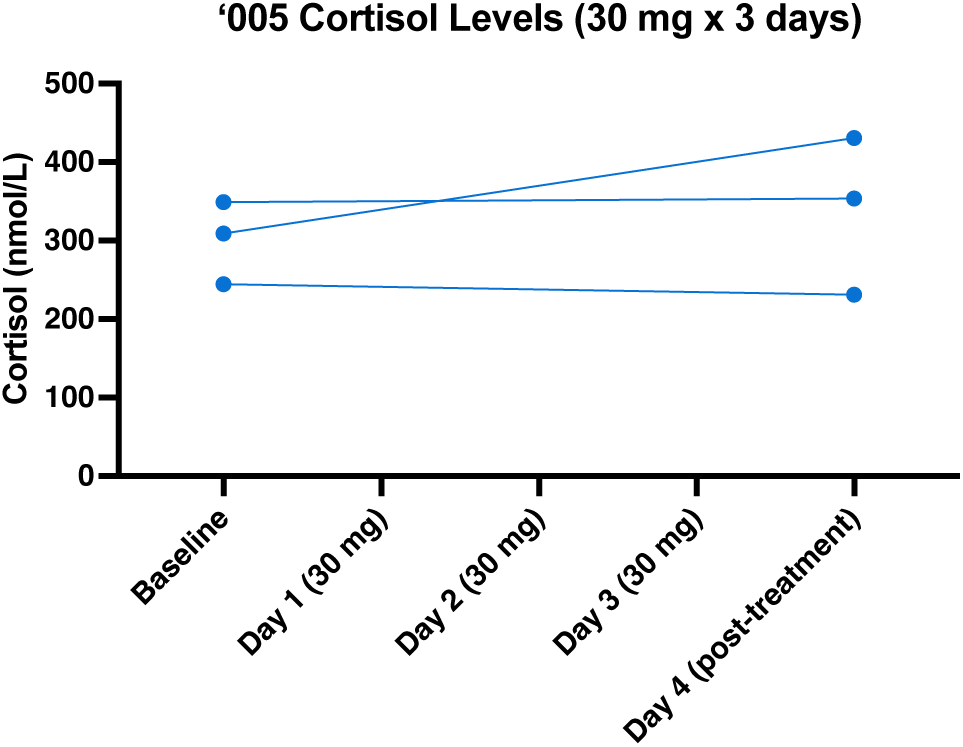
HPA axis preservation in the repeated-dose arm of the first-in-human study (Study 1). Left: Serum cortisol concentrations in the 3 healthy volunteers who received 30 mg IV ’005 once daily for 3 days, at screening and 24 h after the final dose. Cortisol remained within the normal physiologic range in all subjects. Right: ACTH values at the same timepoints confirm absence of pituitary suppression.

#### Study 2 — Dedicated single-/multiple-dose PK and safety study (ICS/LAX/2022-004; ICBio Clinical Research, Bangalore, India)

This single-arm, open-label study enrolled 6 healthy Asian adult male subjects (mean age 34.33 ± 2.66 years; mean BMI 24.17 ± 3.39 kg/m²) who received 30 mg ’005 by IV bolus as a single dose and then once daily for 3 consecutive days, with serial plasma sampling on Day 1 and Day 3 and 24-h urine collection after the first dose. Intact ’005 reached Tmax at 0.5 h on both days and was cleared with a short terminal half-life (1.931 ± 0.383 h on Day 1; 1.959 ± 0.432 h on Day 3), with no evidence of accumulation (Day 3 Cmax 477.167 ± 105.078 vs. Day 1 570.167 ± 55.837 ng/mL). Free dexamethasone was not measurable after the single dose and, after repeated dosing, reached only low concentrations (Day 3 Cmax 1.087 ± 0.339 ng/mL) characterized in the clinical study report as clinically not significant. Intact ’005 was recovered unchanged in urine (Rmax 3.6083 ± 1.28624 ng/mL; AE24 15.0708 ± 5.35986 ng·h/mL; urinary Tmax 2.0 h), supporting renal elimination of the prodrug (Fig. 6). Safety monitoring revealed no deaths, serious adverse events, or clinically significant laboratory, ECG, vital-sign, or glucose changes.

**Fig. 6.**
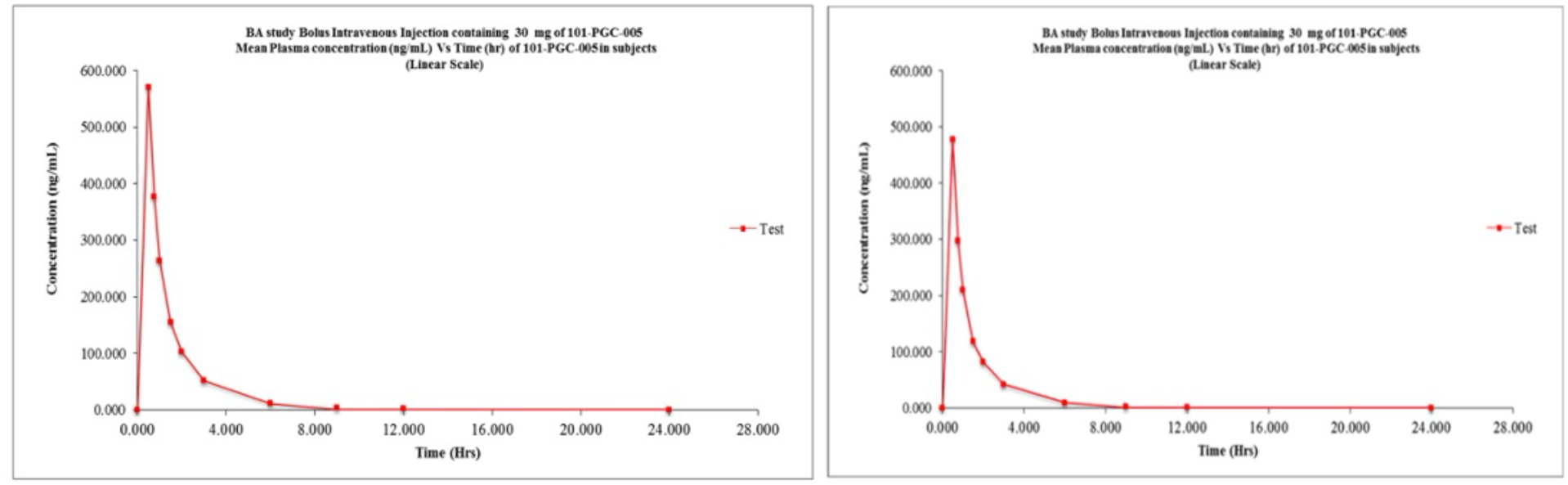
Pharmacokinetics of ’005 in the dedicated single-/multiple-dose study (Study 2). (A) Mean plasma concentrations of ’005 and free dexamethasone following 30 mg IV bolus once daily for 3 days in 6 healthy volunteers, with PK sampling on Days 1 and 3. ’005 reached Tmax at 0.5 h on both days and declined with a terminal half-life of approximately 1.9 h; Day 1 Cmax/AUC0-t were 570.167 ± 55.837 ng/mL and 715.279 ± 107.169 ng·h/mL, and Day 3 values were 477.167 ± 105.078 ng/mL and 577.136 ± 128.457 ng·h/mL. Free dexamethasone was not measurable on Day 1 and appeared only at low, clinically non-significant concentrations after multiple dosing (Day 3 Cmax 1.087 ± 0.339 ng/mL). (B) Urinary excretion of intact ’005 on Day 1 (Rmax 3.6083 ± 1.28624 ng/mL; AE24 15.0708 ± 5.35986 ng·h/mL; Tmax 2.0 h). CSF pharmacokinetics (Study 3) will be reported separately when the clinical study report is finalized.

#### Study 3 — Cerebrospinal fluid PK study (ICS/LAX/2023-002; ICBio Clinical Research, Bangalore, India)

We evaluated whether ’005 remained intact in systemic circulation, limited systemic release of dexamethasone, preserved HPA-axis function, and reached the CSF compartment after intravenous dosing. Across two Phase 1 plasma PK/safety studies and one dedicated CSF PK study in healthy volunteers, ’005 circulated primarily as intact parent prodrug, cleared rapidly from plasma, and produced minimal free-dexamethasone exposure.

In the first-in-human dose-escalation and repeated-dose study (PIF-GC-CP-101; NCT05619770), 15 healthy adult volunteers received single IV doses of 10, 20, or 30 mg ’005, or 30 mg once daily for 3 consecutive days. Free dexamethasone concentrations were very low across cohorts, with a highest individual plasma value of 0.96 ng/mL. In the repeated-dose cohort, morning cortisol and ACTH measured 24 h after the third dose remained within normal physiologic ranges, indicating preservation of HPA-axis function after supratherapeutic repeated dosing (Fig. 5). No dose-limiting toxicities or serious adverse events were observed.

In a dedicated single-/multiple-dose plasma PK and safety study (ICS/LAX/2022-004), 6 healthy adult male subjects received 30 mg ’005 by IV bolus as a single dose and then once daily for 3 consecutive days. Intact ’005 reached peak plasma concentrations rapidly, with median Tmax of 0.5 h on both Day 1 and Day 3, and cleared with a short terminal half-life of 1.931 ± 0.383 h on Day 1 and 1.959 ± 0.432 h on Day 3. There was no evidence of accumulation after repeated dosing. Free dexamethasone was not measurable after the single dose and remained low after repeated dosing, with Day 3 Cmax of 1.087 ± 0.339 ng/mL. Intact ’005 was recovered unchanged in urine, supporting renal elimination of the parent prodrug (Fig. 6 and Table 1). Safety monitoring showed no deaths, serious adverse events, or clinically meaningful changes in laboratory values, ECGs, vital signs, glucose, or injection-site tolerability.

**Table 1.** Dedicated single-/multiple-dose PK and safety study of 30 mg IV ’005 (Study 2; ICS/LAX/2022-004; ICBio, Bangalore, India; n = 6). Values are mean ± SD except Tmax, shown as median (range) as reported. NA = not measurable/not applicable; AE24 = amount/rate of urinary excretion over 24 h as reported. *\*The Day 3 dexamethasone terminal half-life estimate was driven by sparse, very low concentrations and was not interpreted as clinically meaningful systemic dexamethasone exposure in the clinical study report*.

| Analyte / matrix | Day | Cmax / Rmax | AUC0-t / AE24 | Tmax | t1/2 |
| --- | --- | --- | --- | --- | --- |
| 101-PGC-005 / plasma | 1 | 570.167 ± 55.837 ng/mL | 715.279 ± 107.169 ng·h/mL | 0.50 h (0.500-0.500) | 1.931 ± 0.383 h |
| 101-PGC-005 / plasma | 3 | 477.167 ± 105.078 ng/mL | 577.136 ± 128.457 ng·h/mL | 0.500 h (0.500-0.500) | 1.959 ± 0.432 h |
| Dexamethasone / plasma | 1 | NA | NA | NA | NA |
| Dexamethasone / plasma | 3 | 1.087 ± 0.339 ng/mL | 14.134 ± 12.569 ng·h/mL | 0.500 h (0.500-0.750) | 323.296 ± 708.043 h* |
| 101-PGC-005 / urine | 1 | $R_{max}$ 3.6083 ± 1.28624 ng/mL | AE24 15.0708 ± 5.35986 ng·h/mL | 2.000 h (2.000-2.000) | Not reported |

In a dedicated single-dose CSF PK study (ICS/LAX/2023-002), 6 healthy adult male subjects received a 30 mg IV bolus of ’005, with serial plasma and CSF sampling through 24.5 h. A validated LC-MS/MS method quantified ’005 and dexamethasone in human CSF over 0.02–4 ng/mL. Intact ’005 was rapidly measurable in plasma and was also detectable in CSF. In plasma, mean parent-drug Cmax was 654.098 ng/mL, AUC0-inf was 933.874 ng·h/mL, median Tmax was 0.5 h, and terminal half-life was 2.653 h. In CSF, mean parent-drug Cmax was 1.267 ng/mL, AUC0-inf was 8.682 ng·h/mL, median Tmax was 2.185 h (range, 1.820–4.030 h), and terminal half-life was 2.972 h. Plasma exposure to released dexamethasone remained low, with mean Cmax of 0.461 ng/mL and AUC0-24 of 3.998 ng·h/mL.

Single-dose 30 mg ’005 was well tolerated in the CSF PK cohort. No adverse events or serious adverse events were reported, and safety assessments, including vital signs, ECG, hematology, biochemistry, urinalysis, physical examination, and injection-site tolerability, showed no clinically meaningful safety signal. The delayed CSF Tmax relative to plasma, persistence of measurable intact parent drug in CSF, and low circulating free-dexamethasone exposure are consistent with a pharmacologic profile in which central-compartment exposure occurs without clinically meaningful systemic steroid spillover.

Together, these Phase 1 data show that ’005 remains largely intact in systemic circulation, clears rapidly from plasma, is excreted unchanged in urine, produces absent or very low systemic free-dexamethasone exposure, preserves cortisol and ACTH after repeated supratherapeutic dosing, and reaches the CSF compartment after IV administration. These findings support the pharmacologic premise of macrophage-targeted glucocorticoid delivery with limited systemic dexamethasone exposure and preserved endocrine physiology.

### Adaptive Phase II/III Randomized Controlled Trial: Study Design

A prospective, randomized, open-label, active-controlled adaptive Phase II/III trial (Protocol ICS/LAX/2022-005; ClinicalTrials.gov NCT05656521) was conducted at six sites across India (Pune ×2, Mumbai, Nellore, Bengaluru, Gandhinagar) under approval from the Central Drugs Standard Control Organisation (CDSCO), with independent Ethics Committee review at each site and oversight of the Phase II interim by an independent Data Safety Monitoring Board (DSMB). The study was co-sponsored by 101 Therapeutics and Laxai Life Sciences, with Insignia Clinical Services (Delhi) as CRO.

The adaptive design enrolled an initial Phase II cohort of 62 patients (31 per arm) at four sites from January to April 2023. Following DSMB interim review of unblinded Phase II data, continuation to Phase III was approved. Phase III enrolled an additional 247 patients (124 vs. 123) across the same four original sites plus five new sites from January 2024 to July 2025, capturing a distinct COVID-19 variant landscape and higher/mixed vaccination background relative to the Phase II cohort. This temporal and epidemiological separation constitutes a prospective internal replication: the original four sites reproduced their own findings in a different pathogen environment, while five independent sites provided external confirmation. Participant flow through the adaptive trial is shown in Fig. 11.

Eligible patients were adults hospitalized with moderate COVID-19 (WHO ordinal scale score 4–5, supplemental oxygen requirement without mechanical ventilation). No restrictions were placed on comorbidities or concomitant medications (excluding prior corticosteroids or immunosuppressants within 14 days). Participants were randomized 1:1 to receive either ’005 20 mg IV bolus once daily for 3 days plus standard of care (SOC), or dexamethasone 6 mg IV once daily for a minimum of 5 and maximum of 10 days (physician-determined) plus SOC. The protocol explicitly permitted investigators to supplement with any additional medication, including corticosteroids, at any time if clinical judgment deemed the assigned treatment insufficient.

### Demographic and disease characteristics at baseline.**\***

| Characteristic | '005 + SOC (N = 155) | Dexamethasone + SOC (N = 154) | Total (N = 309) |
| --- | --- | --- | --- |
| **Demographic characteristics** |  |  |  |
| Age — yr | 41.1 ± 11.0 | 43.2 ± 11.0 | 42.1 ± 11.0 |
| Female sex — no. (%) | 31 (20.0) | 28 (18.2) | 59 (19.1) |
| Male sex — no. (%) | 124 (80.0) | 126 (81.8) | 250 (80.9) |
| Height — cm | 165.9 ± 6.4 | 166.5 ± 6.5 | 166.2 ± 6.5 |
| Weight — kg | 67.4 ± 8.4 | 67.1 ± 7.9 | 67.2 ± 8.2 |
| Body-mass index† | 24.5 ± 2.9 | 24.2 ± 3.0 | 24.4 ± 2.9 |
| **Body-mass index category — no. (%)** |  |  |  |
| <25 | 93 (60.0) | 91 (59.1) | 184 (59.5) |
| ≥25 | 62 (40.0) | 63 (40.9) | 125 (40.5) |
| **Race or ethnic group — no. (%)‡** |  |  |  |
| Asian | 155 (100) | 154 (100) | 309 (100) |
| **COVID-19 vaccination status — no. (%)** |  |  |  |
| Yes | 75 (48.4) | 66 (42.9) | 141 (45.6) |
| No | 80 (51.6) | 88 (57.1) | 168 (54.4) |
| **Medical history — no. (%)** |  |  |  |
| Any medical history | 44 (28.4) | 50 (32.5) | 98 (31.7) |
| Type 2 diabetes mellitus | 25 (16.1) | 30 (19.5) | 57 (18.4) |
| Hypertension | 15 (9.7) | 24 (15.6) | 39 (12.6) |
| Anemia | 6 (3.9) | 5 (3.2) | 13 (4.2) |
| **Baseline COVID-19 disease characteristics** |  |  |  |
| WHO 11-point ordinal-scale score — no. (%) |  |  |  |
| 4 | 80 (51.6) | 74 (48.1) | 154 (49.8) |
| 5 | 75 (48.4) | 80 (51.9) | 155 (50.2) |
| Supplemental oxygen at baseline — no. (%) | 75 (48.4) | 80 (52.0) | 155 (50.2) |
| Oxygen saturation — % | 91.9 ± 0.7 | 91.9 ± 0.8 | — |
| Temperature — °F | 100.9 ± 1.4 | 101.1 ± 1.2 | — |
| Pulse rate — beats/min | 84.2 ± 6.7 | 83.6 ± 6.0 | — |
| Lung-injury score§ | 2.0 ± 0.3 | 2.0 ± 0.2 | 2.0 ± 0.2 |

| **Inflammatory and coagulation biomarkers** |  |  |  |
| --- | --- | --- | --- |
| C-reactive protein — mg/L | 29.0 ± 33.8 | 25.7 ± 20.4 | — |
| D-dimer — ng/mL¶ | 682.9 ± 583.3 | 1110.5 ± 4340.0 | — |
| Ferritin — ng/mL | 251.0 ± 205.9 | 240.1 ± 208.2 | — |
| Interleukin-6 — pg/mL¶ | 16.9 ± 25.2 | 15.5 ± 16.1 | — |
| Neutrophil-to-lymphocyte ratio¶ | 2.83 ± 2.60 | 3.09 ± 3.18 | — |
| Tumor necrosis factor-α — pg/mL¶ | 59.5 ± 107.9 | 56.3 ± 72.8 | — |

\* Plus-minus values are means ± SD. Percentages are based on the safety analysis set unless otherwise indicated. SOC denotes standard of care.
† The body-mass index is the weight in kilograms divided by the square of the height in meters.
‡ All enrolled participants were treated at sites in India and were reported as Asian.
§ Lung-injury score data were available for 155 patients in the '005 + SOC group and 153 patients in the dexamethasone + SOC group.
¶ Baseline D-dimer data were available for 154 and 153 patients, interleukin-6 data for 154 and 152 patients, neutrophil-to-lymphocyte ratio data for 154 and 152 patients, and tumor necrosis factor-α data for 152 and 147 patients in the '005 + SOC and dexamethasone + SOC groups, respectively.

### Phase II/III Primary Endpoint: Clinical Improvement

The primary endpoint was time to ≥2-point improvement on the WHO 11-point ordinal scale, analyzed by Cox proportional hazards model (non-inferiority margin 0.70). Across all 309 randomized patients (ITT = Full Analysis Set = per-protocol; zero dropouts), ’005 was superior to dexamethasone: HR 2.31 (95% CI 1.83–2.93; p < 0.0001; Fig. 7). Median time to improvement was 3 days for ’005 versus 4 days for dexamethasone. The hazard ratio was consistent with that observed in the Phase II cohort alone (HR 2.31), indicating that the effect was reproduced exactly across the two-phase adaptive design.

**Fig. 7.**
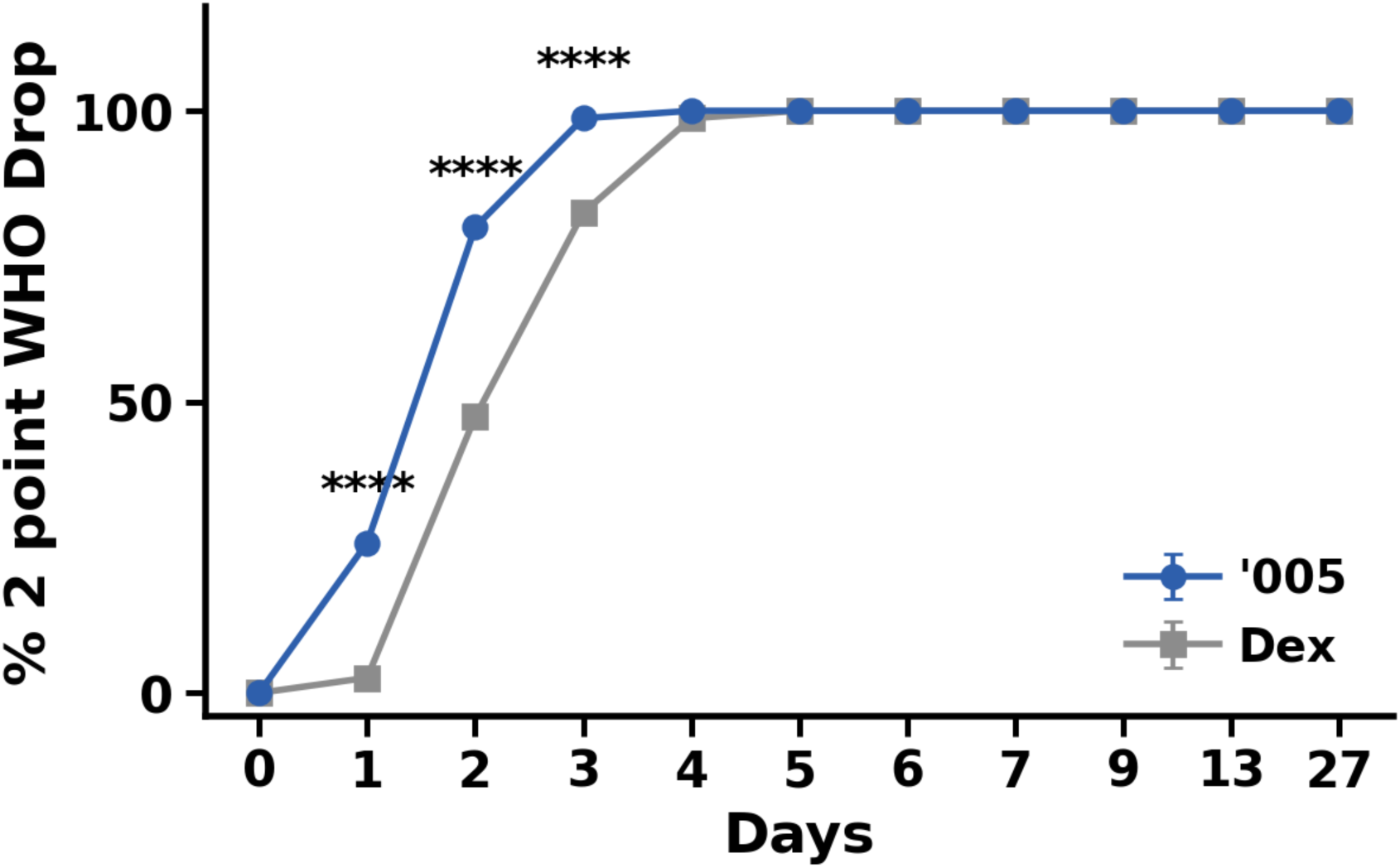
Time to ≥2-point WHO ordinal-scale improvement, all 309 randomized patients. Cumulative percentage of patients achieving a ≥2-point improvement from baseline on the WHO 11-point ordinal scale, plotted against days since baseline, for ’005 + SOC (blue, n = 155) and dexamethasone + SOC (grey, n = 154). ’005 was superior on the primary endpoint (time to ≥2-point improvement) by Cox proportional-hazards analysis: HR 2.31 (95% CI 1.83–2.93), p < 0.0001 against a non-inferiority margin of 0.70; median time to improvement was 3 days for ’005 versus 4 days for dexamethasone. The cumulative proportion improved was significantly higher with ’005 on days 1, 2 and 3 (25.8% vs 2.6%, 80.0% vs 47.4%, and 98.7% vs 82.5%, respectively), with both arms reaching ∼100% by days 4–5. Asterisks denote between-arm comparisons at each day by Fisher’s exact test (****p < 0.0001); days without asterisks were not significant. Analysis population was ITT = full analysis set = per-protocol (zero dropouts).

**Fig. 8.**
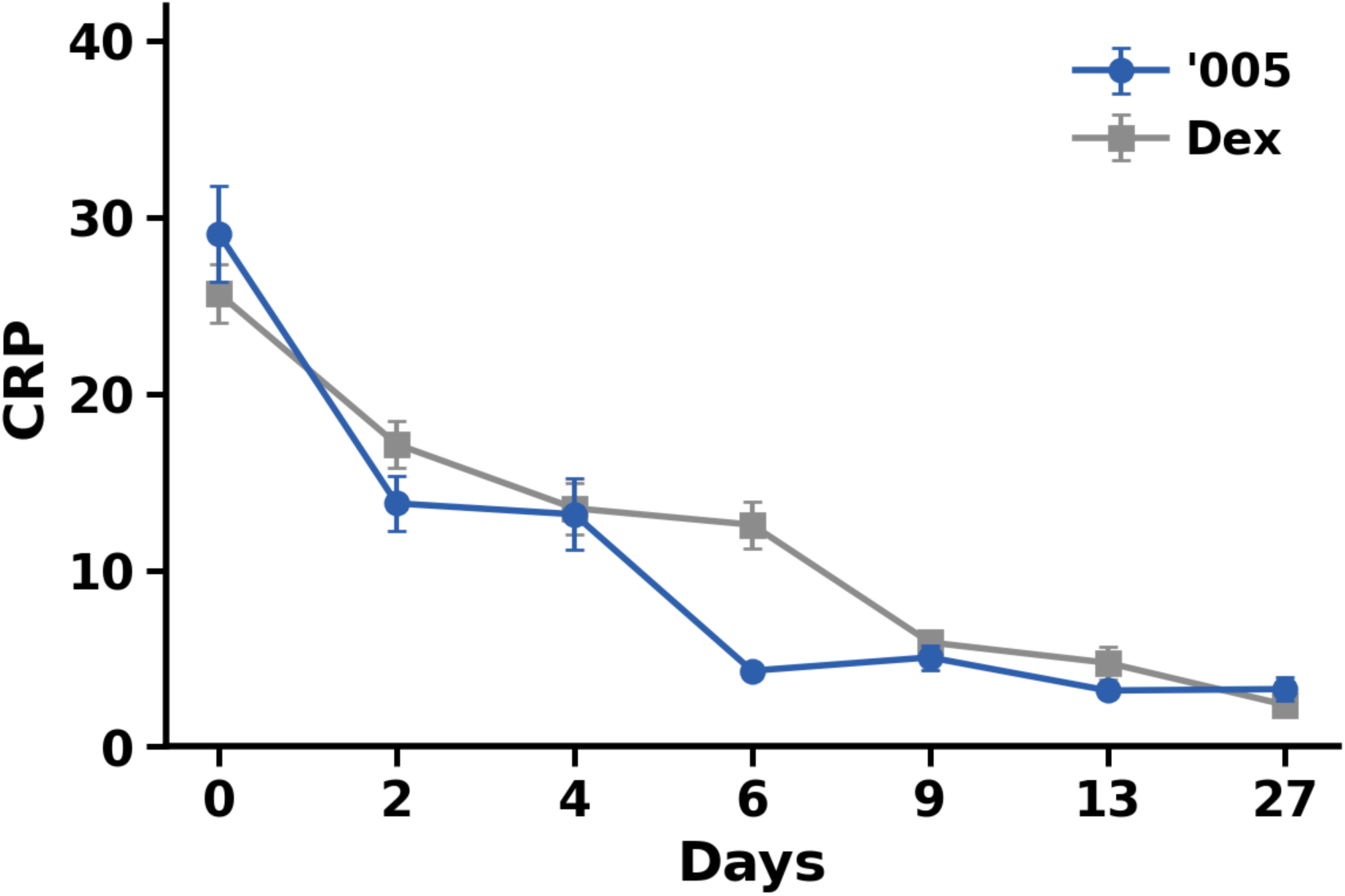
C-reactive protein over time. Mean serum CRP (mg/L) by days since baseline for ’005 + SOC (blue) and dexamethasone + SOC (grey); error bars denote SEM. Baseline CRP was comparable between arms (29.1 ± 2.7 mg/L, n = 155 vs 25.7 ± 1.6 mg/L, n = 154). CRP fell faster in the ’005 arm from day 2 onward (13.8 ± 1.6 vs 17.1 ± 1.4 mg/L) and remained numerically lower through the in-hospital period, converging by the day 9–27 follow-up. No individual visit-day comparison reached statistical significance by Mann–Whitney U test (closest: day 2, p = 0.09; day 9, p = 0.06), so no significance asterisks are shown. The apparent widening at day 6 (4.3 ± 0.4 vs 12.6 ± 1.3 mg/L) reflects sparse on-treatment sampling at that visit (n = 5 vs 40) and should be interpreted with caution. Sample size declines at later in-hospital days as patients were discharged.

Day-by-day cumulative improvement rates are shown in Table 2: A critical confirmatory finding was the zero rate of investigator-initiated corticosteroid supplementation in the ’005 arm. Despite the protocol explicitly granting physicians the right to add any medication, including systemic corticosteroids, if they judged the patient’s response insufficient, no investigator exercised this right in any of the 155 patients receiving ’005 across 6 centers over 2.5 years. This constitutes a silent, unbiased clinical endorsement: physicians with full discretionary authority and real-world clinical responsibility, observing patients in real time, never concluded that ’005 was inadequate. Comparator-arm patients received a mean cumulative dexamethasone dose of approximately 30–36 mg over the observation period.

**Table 2.** Day-by-day cumulative rates of ≥2-point WHO ordinal scale improvement. All 309 randomized patients analyzed (ITT = per-protocol). pp = percentage points; NS = not significant at Day 4 due to floor effect (both arms approaching 100%).

| Day | '005 + SOC | Dexa + SOC | Δ (percentage points) | p-value |
| --- | --- | --- | --- | --- |
| Day 1 | 25.81% (40/155) | 2.60% (4/154) | +23.2 pp | < 0.0001 |
| Day 2 | 80.00% (124/155) | 47.40% (73/154) | +32.6 pp | < 0.0001 |
| Day 3 | 98.71% (153/155) | 82.5% (127/154) | +16.2 pp | < 0.0001 |
| Day 4 | 100% (155/155) | 98.70% (152/154) | +1.3 pp | NS |

### Phase II/III Secondary Endpoints

Viral clearance: ’005 significantly accelerated SARS-CoV-2 PCR negativity (HR 1.47; 95% CI 1.17–1.84; p = 0.0001), with median clearance at 3 days versus 5 days in the dexamethasone arm. This is consistent with the CXCL10 preservation mechanism observed in vitro: by retaining interferon-pathway signaling, ’005 does not impair the antiviral immune response that drives viral elimination.

Hospital discharge: Cumulative discharge rates were significantly higher in the ’005 arm from Day 3 onward. By Day 5, 77.4% of ’005 patients had been discharged versus 29.0% of dexamethasone patients (p = 0.0023). By Day 6, 87.1% versus 41.9% (p = 0.0023). All patients in both arms were discharged by Day 8. The magnitude of early discharge difference, 48 percentage points at Day 5, has direct health-economic implications, as each bed-day avoided reduces hospital cost and infection transmission risk.

Oxygenation recovered significantly faster with ’005 on Days 1–4, with convergence by Day 5 onward (Fig. 10). Fever resolution was similarly faster in the ’005 arm on Days 1–4, consistent with more rapid macrophage debulking and cytokine burden reduction (Fig. 9).

**Fig. 9.**
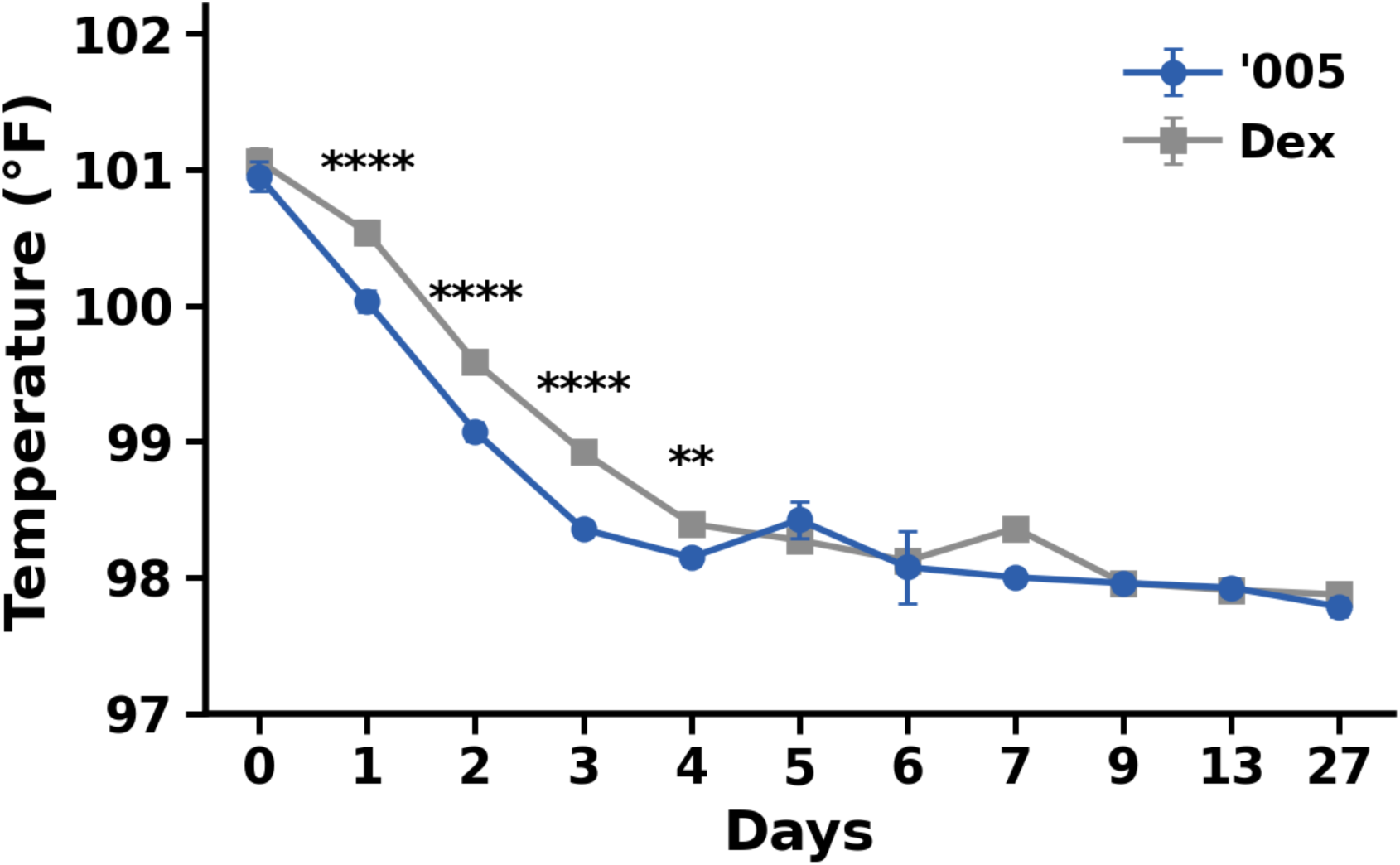
Defervescence. Mean body temperature (°F) by days since baseline for ’005 + SOC (blue) and dexamethasone + SOC (grey); error bars denote SEM; n = 155 (’005) and 154 (dexamethasone) at baseline, declining at later in-hospital visits as patients were discharged. Baseline temperatures were comparable (100.9 ± 0.1 vs 101.1 ± 0.1 °F) and both arms were afebrile by approximately day 4. Temperature normalized significantly faster in the ’005 arm on days 1–4 (day 1, 100.03 ± 0.08 vs 100.54 ± 0.09; day 2, 99.07 ± 0.07 vs 99.59 ± 0.08; day 3, 98.35 ± 0.06 vs 98.92 ± 0.07 °F), with no between-arm difference thereafter. Asterisks denote between-arm comparisons at each day by Welch’s t-test (**p < 0.01, ****p < 0.0001); days without asterisks were not significant.

**Fig. 10.**
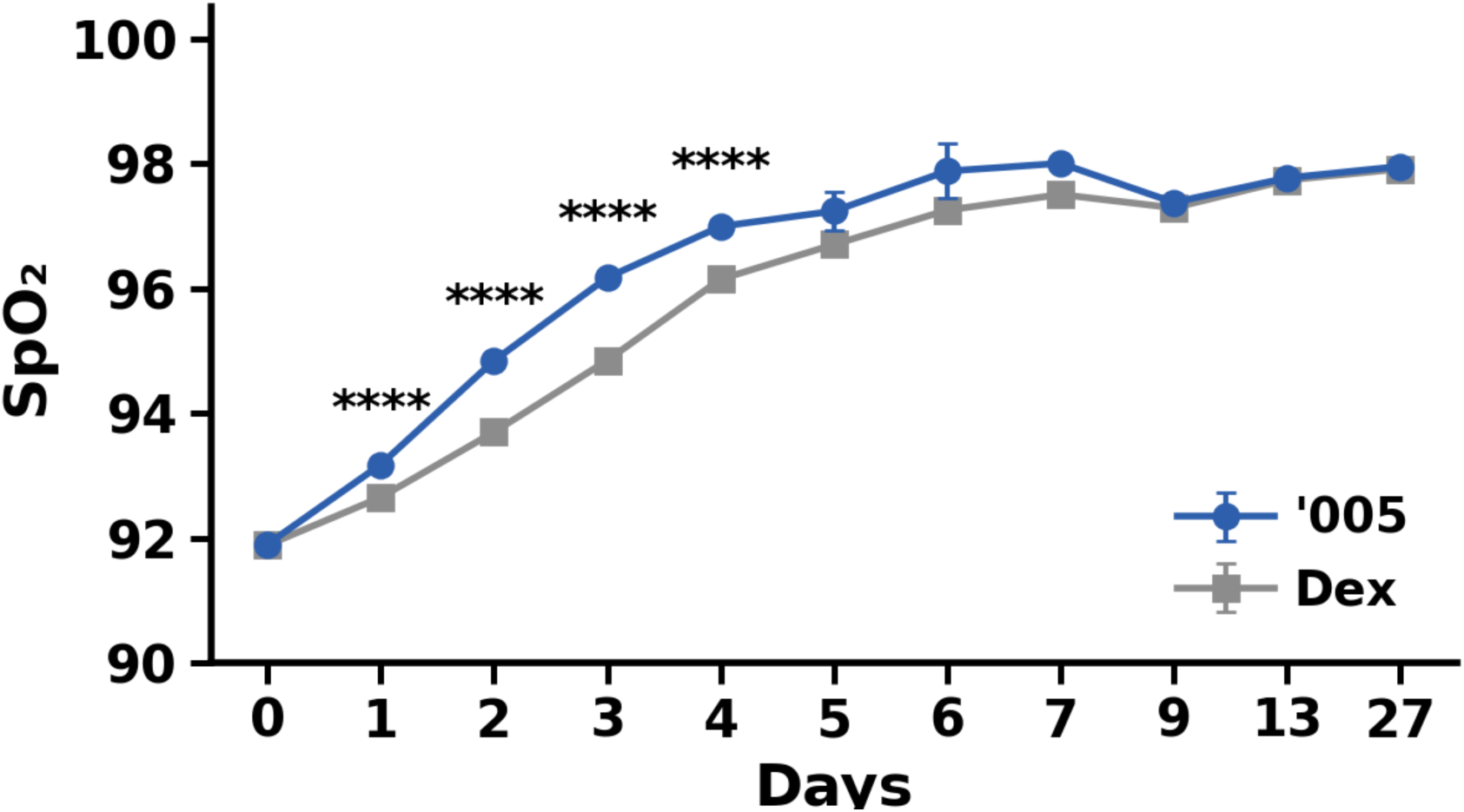
Oxygenation recovery. Mean oxygen saturation (SpO₂, %) by days since baseline for ’005 + SOC (blue) and dexamethasone + SOC (grey); error bars denote SEM; n = 155 (’005) and 154 (dexamethasone) at baseline, declining at later in-hospital visits as patients were discharged. Baseline SpO₂ was comparable (91.9 ± 0.1% in both arms). Oxygenation recovered significantly faster with ’005 on days 1–4 (day 2, 94.8 ± 0.1 vs 93.7 ± 0.1; day 3, 96.2 ± 0.1 vs 94.8 ± 0.1; day 4, 97.0 ± 0.1 vs 96.1 ± 0.1%), converging by day 5 onward. Asterisks denote between-arm comparisons at each day by Welch’s t-test (****p < 0.0001); days without asterisks were not significant.

**Fig. 11.**
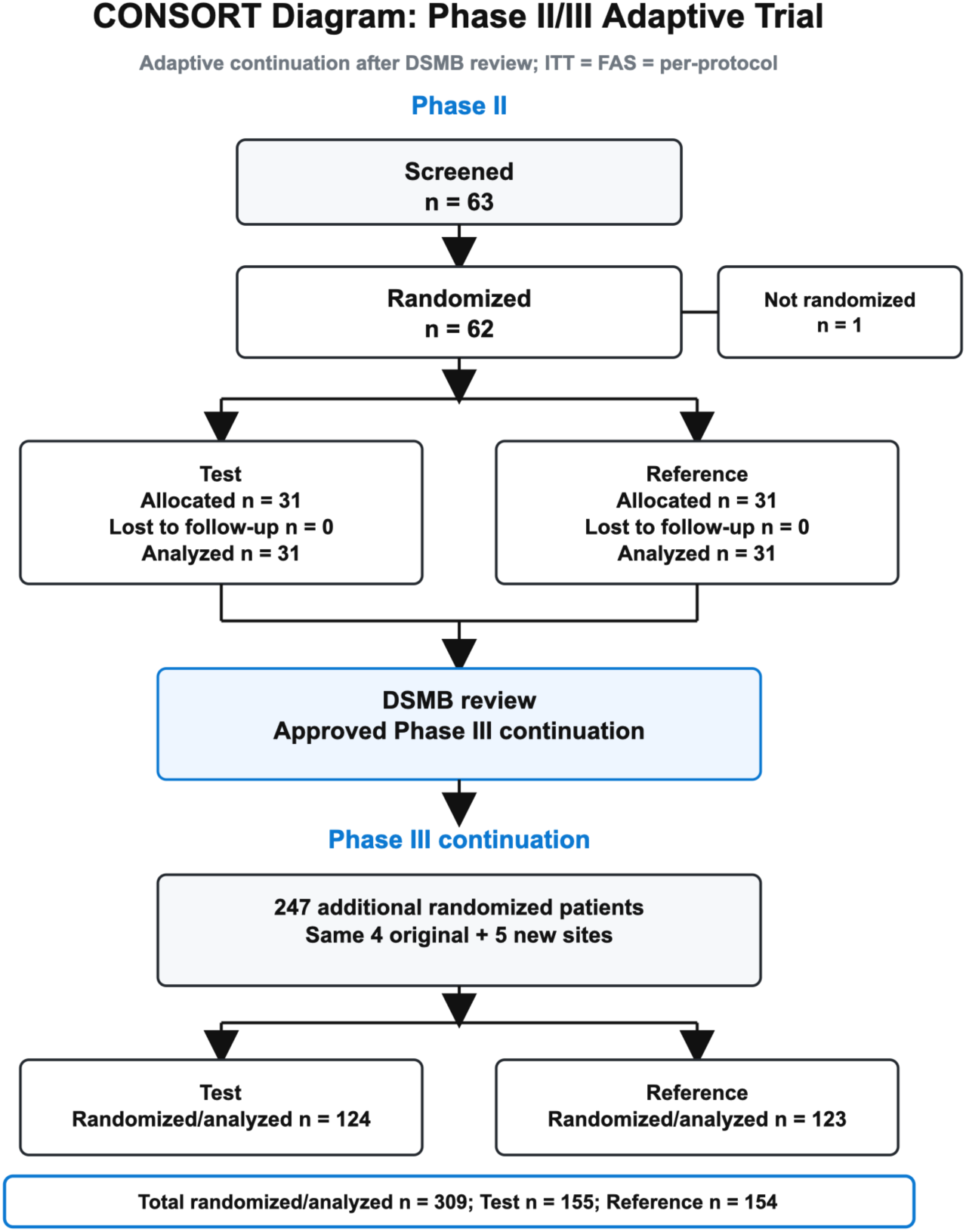
CONSORT diagram of the adaptive Phase II/III trial. Participant flow through the adaptive design. Phase II: 63 screened, 62 randomized 1:1 (’005 n = 31; dexamethasone n = 31), 1 not randomized, 0 lost to follow-up, and all randomized participants analyzed. After the independent Data Safety Monitoring Board reviewed unblinded Phase II data, continuation to Phase III was approved. Phase III: 247 additional patients randomized (’005 n = 124; dexamethasone n = 123) across the 4 original plus 5 new sites. In total, 309 patients were randomized and 309 analyzed (’005 n = 155; dexamethasone n = 154); ITT = full analysis set = per-protocol, with zero dropouts.

Biomarkers: CRP declined numerically faster in the ’005 arm from Day 2 onward and remained numerically lower through the in-hospital period, but no individual visit-day comparison reached statistical significance (Fig. 8). IL-6 and D-dimer showed consistent directional superiority for ’005. HRCT lung improvement at Day 28 was equivalent between arms (LS mean difference −0.05; p = 0.376), consistent with equivalent long-term respiratory recovery despite faster early clinical improvement. Zero ICU admissions and zero deaths occurred in either arm.

### Safety

TEAEs occurred in 54.84% of ’005 patients and 54.55% of dexamethasone patients (p = 0.958), establishing equivalence of adverse event rates at approximately five times the sample size of the Phase II cohort (Table 3). The Phase II cohort showed numerically higher TEAEs in the ’005 arm (35.5% vs. 19.4%), likely reflecting more granular adverse event capture in a smaller, intensively monitored early cohort; the larger Phase III population demonstrated rate convergence.

**Table 3.** Safety summary, Phase II/III combined (n = 309). TEAE = treatment-emergent adverse event; SAE = serious adverse event; SUSAR = suspected unexpected serious adverse reaction; GC = glucocorticoid.

| Parameter | '005 + SOC (n=155) | Dexa + SOC (n=154) | p-value |
| --- | --- | --- | --- |
| Any TEAE, n (%) | 85 (54.84%) | 84 (54.55%) | 0.958 |
| Drug-related TEAE, n | 17 | 16 | 0.687 |
| Grade $\geq 3$ events | 0 | 0 | — |
| SAEs | 0 | 0 | — |
| Deaths | 0 | 0 | — |
| Discontinuations due to AE | 0 | 0 | — |
| SUSARs | 0 | 0 | — |
| GC add-on (investigator-initiated) | 0 | N/A | — |
| Cumulative exogenous corticosteroid | 0 mg | ~30–36 mg Dex | — |

All TEAEs were Grade 1 (mild) or Grade 2 (moderate); no Grade 3 or higher events occurred in either arm. Drug-related TEAEs were 17 in the ’005 arm versus 16 in the dexamethasone arm (p = 0.687). No serious adverse events, deaths, SUSARs, or discontinuations were recorded in either arm. No patient received a corticosteroid add-on in the ’005 arm. The ’005 arm received zero milligrams of exogenous corticosteroid versus approximately 30–36 mg cumulative dexamethasone in the comparator arm, establishing that equivalent safety outcomes were achieved with complete steroid sparing.

## Discussion

This work provides a multilevel, independently replicated evidence base for a new class of macrophage-targeted corticosteroids. Across mechanistic assays, pathogen-diverse preclinical models, two completed human PK studies (with a cerebrospinal fluid study underway), and an adaptive Phase II/III trial in 309 patients powered for non-inferiority but achieving superiority, ’005 produced rapid and durable clinical benefit, preserved antiviral immunity, and avoided the systemic endocrine and immunosuppressive toxicities that have constrained GC use since the 1940s.

The completed human PK studies collectively address whether ’005 releases dexamethasone systemically at clinically meaningful levels; the answer is no. In the first-in-human Israeli study (Study 1), free plasma dexamethasone was very low across single- and repeated-dose cohorts (peak 0.96 ng/mL), and morning cortisol and ACTH remained within normal ranges after three consecutive days of 30 mg dosing, the clearest in vivo test of systemic GC exposure, since even low systemic dexamethasone can suppress ACTH within hours. The dedicated Indian PK study (Study 2) confirmed rapid clearance (Tmax ∼0.5 h; half-life ∼1.9 h), absence of accumulation, no measurable free dexamethasone after single dosing and only low, clinically non-significant concentrations after repeated dosing, and renal excretion of intact prodrug. A dedicated CSF PK study (Study 3) is evaluating whether ’005 reaches CNS-adjacent compartments, building on preclinical evidence that ’005, but not free dexamethasone, crosses the blood–brain barrier. Together, these data indicate that the oxime-ether linker is sufficiently stable in systemic circulation to avoid clinically meaningful free dexamethasone exposure, with pharmacologic glucocorticoid activity concentrated away from the systemic endocrine axis.

The Phase II/III trial design deserves specific attention for its evidentiary quality. The non-inferiority-powered adaptive design, with prospective DSMB gating between Phase II and III, is a conservative framework: the fact that the study achieved superiority on the primary endpoint means the observed treatment effect substantially exceeded the minimum clinically meaningful threshold. The hazard ratio of 2.31 was identical in the 62-patient Phase II cohort and the 309-patient combined dataset, an internal replication that eliminates the possibility of a chance finding in either cohort. The temporal separation of the two phases, spanning over one year and different COVID-19 variant and vaccination landscapes, further strengthens the inference: the mechanism works independently of specific pathogen variants or baseline immune status.

The zero rate of investigator-initiated corticosteroid supplementation deserves emphasis as a clinical quality metric that is not captured in standard efficacy analyses. Physicians observing individual patients in real time, with complete clinical authority, never judged ’005 to be inadequate for 155 consecutive patients across 6 hospitals and 2.5 years. This is not a statistical outcome, it is a practitioner judgment expressed 155 times without exception. It represents a different kind of evidence than a hazard ratio: it is the signal that practicing physicians would reach for ’005 again.

The superiority on viral clearance provides mechanistic confirmation in patients. The CXCL10 data from infected macrophages showed that ’005 suppresses IL-6 and TNF to dexamethasone-equivalent levels while preserving interferon-pathway antiviral signaling. The accelerated PCR negativity in patients, median 3 versus 5 days, HR 1.47, is the clinical expression of that preserved antiviral competence.

Dexamethasone, by broadly suppressing GR signaling including in lymphocytes and other immune cells, carries a theoretical risk of impaired viral clearance; ’005, by confining its GC activity to macrophages while preserving the broader immune response, may be intrinsically better suited to infectious inflammatory settings.

The hospital discharge data carry direct clinical and health-systems implications. A 48-percentage-point difference in discharge rates at Day 5 (77.4% vs. 29.0%) translates to meaningful reduction in bed-days, hospital-acquired infection exposure, and demand on inpatient resources. The safety equivalence data, no Grade 3+ events, no SAEs, equivalent TEAE rates in the larger Phase III population, indicate that this earlier discharge was not achieved at the cost of undertreated disease or premature release but reflected genuine, faster resolution of the underlying inflammatory process.

Conventional GCs remain the fastest and most reliable anti-inflammatory agents in acute medicine, yet systemic exposure suppresses the HPA axis, blunts adaptive immunity, and drives metabolic, skeletal, and neuropsychiatric toxicities [11,23]. The central innovation here is to retain pharmacologic potency while relocating exposure: ’005 accumulates inside CD206⁺ macrophages via low-affinity, recycling-competent mannose receptor engagement, then releases active dexamethasone intracellularly to trigger apoptosis and transcriptional reprogramming of pathogenic macrophage subsets. By confining the bulk of GC action to the inflammatory compartment, ’005 achieves rapid, multi-pathway cytokine suppression without the endocrine and immunosuppressive consequences.

A mechanistic property of ’005 that has important therapeutic implications deserves explicit attention: ’005 depletes pathogenic activated macrophages but does not block the activation of newly differentiated macrophages arising from the circulating monocyte pool. The monocyte compartment is effectively inexhaustible under physiologic conditions, continuously producing naïve macrophage precursors that, once tissue-resident and under HPA control, adopt homeostatic rather than pathogenic phenotypes. By removing the existing burden of dysregulated, cortisol-resistant macrophages while leaving the renewal pathway intact, ’005 enables a functional reset: the depleted pathogenic population is replaced by HPA-responsive macrophages capable of executing normal resolution and repair programs. This is qualitatively different from the action of cytokine inhibitors or Janus kinase (JAK) inhibitors, which suppress signaling across both pathogenic and homeostatic macrophage populations without addressing the underlying cellular burden. Equally important, ’005 does not neutralize or bind soluble cytokines and chemokines already released into the extracellular space. The upstream signals that drove macrophage activation, DAMPs, pathogen-associated molecular patterns, and tissue-derived activating stimuli, remain present and available to guide the incoming population of monocyte-derived macrophages through normal, time-limited activation and resolution sequences. Rather than severing the signaling environment that macrophages require for tissue surveillance and repair, ’005 simply removes the cells that have become irreversibly dysregulated within it.

The bone density finding at 30 mg/kg in the GLP toxicology study warrants specific mechanistic comment. CD206+ macrophages include the osteal macrophage (osteomac) subpopulation that resides on bone surfaces and contributes to bone remodeling homeostasis by supporting osteoblast activity and modulating osteoclastogenesis via the RANKL/OPG axis. F4/80+CD206+ M2-like macrophages in arthritic synovium have been shown to express RANK and differentiate into functional osteoclasts via RANKL signaling, establishing CD206+ macrophages as osteoclast precursors in certain contexts. At 30 mg/kg administered daily for 28 days, approximately 105× the human mg/kg dose (20 mg flat IV in a 70 kg patient) and 9× the number of clinical doses, sustained depletion of the CD206+ osteal population appears sufficient to perturb normal macrophage-mediated bone turnover regulation, producing the observed Marked findings in 4 of 6 animals. We interpret this as an exaggerated pharmacological effect at extreme supratherapeutic exposure rather than an adverse toxicological finding in the conventional sense: it is produced by the same CD206-targeting mechanism that drives therapeutic action, is predicted by known osteal macrophage biology, and is entirely distinct from the osteoblast suppression and calcium dysregulation that characterize conventional glucocorticoid bone toxicity. The NOAEL is accordingly maintained at 30 mg/kg. The 20 mg/kg dose, itself approximately 70× the human mg/kg equivalent, produced no bone density changes across 28 daily doses, providing a wide margin of preclinical safety relative to the clinical regimen of 20 mg flat IV for 3 days total. Prospective monitoring of bone turnover markers is nonetheless warranted in future chronic-indication studies to characterize the relationship between dosing interval and CD206+ osteal macrophage recovery kinetics.

Biologic cytokine inhibitors and JAK inhibitors have transformed chronic disease management, but their kinetics and scope are mismatched to acute systemic inflammation [25,26]. They manifest therapeutic effects only over days to weeks, incompletely suppress inflammatory networks, and can destabilize adrenal regulation by altering cytokine feedback [26, 27]. In contrast, ’005 rapidly debulks the cellular engine of the cytokine program, activated macrophages, while sparing monocytes and preserving antiviral CXCL10. The net effect is fast resolution of inflammatory tone with minimal collateral immunosuppression.

A defining and underappreciated feature of ’005 is not merely that it preserves HPA axis function, it is that it actively leverages the HPA axis as an efficacy partner. Endogenous cortisol is released in precisely calibrated circadian and ultradian pulses that respond to the actual inflammatory signal in real time. Unlike any exogenous GC, it crosses the blood-brain barrier efficiently, acts on GR-expressing cells across every tissue compartment, lymphocytes, endothelial cells, neurons, newly differentiating macrophages from the monocyte pool, and governs the systemic coordination of immune resolution, vascular tone, neuroinflammation prevention, and tissue repair. This pulsatile, demand-responsive, ubiquitous anti-inflammatory intelligence cannot be replicated by any pharmaceutical; it can only be preserved or destroyed. Systemic dexamethasone destroys it by overriding the HPA axis with a flat, non-pulsatile exogenous signal that triggers GR downregulation, adrenal suppression, and the maladaptive cycle of GC resistance. ’005 preserves it, and more than preserves it: by depleting the chronically activated, IL-6/TNF-producing CD206⁺ macrophages that were driving DAMPs and cytokines at levels that overwhelmed HPA control, ’005 removes the inflammatory burden that was dysregulating cortisol release in the first place. The result is a restoration of the condition for physiologic cortisol pulsatility to resume, allowing endogenous cortisol to govern the resolution of inflammation in newly recruited macrophages, lymphocytes, CNS parenchymal cells, and endothelial tissues that ’005 cannot directly access. ’005 performs the cellular heavy lifting, depleting the GC-resistant, HPA-uncoupled pathogenic macrophage burden, and cortisol completes the resolution, acting on every other cell type with the precision that eight decades of pharmacology have failed to replicate artificially. The two mechanisms are genuinely complementary: neither achieves alone what they achieve together.

By debulking acute macrophage inflammation and preserving cortisol rhythms, ’005 may prevent the drift toward chronic, multisystem disease. The macrophage-depletion arm of its mechanism suggests potential in atherosclerosis, peripheral artery disease, metabolic dysfunction-associated steatohepatitis, long COVID, neurodegeneration (amyloid/tau-laden microglia), tumor-associated macrophage-rich tumors, and age-related macular degeneration, where long-lived CD206⁺ macrophages sustain pathology [15, 28].

Our study has limitations. The Phase II/III trial was conducted entirely in India; while GCP compliance, multi-site design, and temporal replication substantially strengthen generalizability, US and European regulatory submissions will require complementary studies in those populations. The non-inferiority design, though conservative, means the primary analysis was powered to detect equivalence rather than superiority; post hoc superiority interpretations should be considered hypothesis-generating for the formal Phase III programs they will inform. The HPA preservation data derive from healthy volunteers at therapeutic doses; the endocrine response in critically ill patients, where baseline HPA dysregulation is common, will require prospective characterization. Finally, the ’resistance-proof’ claim is mechanistic, longitudinal studies are needed to quantify resistance propensity under repeated or sustained exposures.

Future development should test five paths. First, acute indications with high endocrine vulnerability (sepsis, severe influenza, systemic cytokine-release states), powered for time-to-recovery and organ-support-free days, where the pathogen-agnostic, HPA-preserving mechanism of ’005 is directly applicable. Second, chronic inflammatory diseases with endocrine dysregulation (rheumatoid Arthritis, irritable bowl syndrome, systemic idiopathic juvenile arthritis flares), to evaluate whether short ’005 pulses reduce flare duration while improving cortisol rhythms and lowering cumulative immunosuppression relative to biologics and JAK inhibitors.

Third, cytokine release syndrome (CRS) and immune effector cell-associated neurotoxicity syndrome (ICANS) arising from cancer immunotherapy, including CAR-T cell infusion, bispecific antibody step-up dosing, and dual checkpoint inhibitor regimens. Current management of grade 2–4 CRS relies on IL-6 blockade (tocilizumab) and systemic dexamethasone, both of which carry a risk of impairing the very T-cell and NK-cell effector activity that defines therapeutic efficacy. ’005 offers a mechanistically distinct approach: by targeting CD206⁺ macrophages, the primary producers of IL-6, IL-1, and TNF in CRS, while sparing T cells, NK cells, and monocytes entirely, ’005 may allow rapid cytokine debulking without blunting the antitumor immune response. The preservation of CXCL10 and the selectivity for macrophages over lymphocytes, demonstrated in our in vitro data, provide direct mechanistic support for this hypothesis. Prospective evaluation of ’005 as a CRS intervention, either replacing or reducing systemic dexamethasone in immunotherapy protocols, represents an urgent and scientifically tractable next step.

Fourth, the intersection of visceral adipose inflammation and contemporary rapid weight loss interventions,including GLP-1/GIP receptor agonists (incretins) and bariatric surgery. Both approaches achieve profound, rapid reductions in adipose mass, but the kinetics of adipose tissue remodeling are mismatched to the kinetics of weight loss. Visceral adipose tissue in chronic obesity is densely infiltrated by long-lived, chronically activated CD206⁺ macrophages that drive a sustained local and systemic inflammatory state [13]. During rapid fat mobilization, dying adipocytes release lipid-laden contents and danger signals that amplify macrophage activation and can trigger a transient but pronounced inflammatory surge, contributing to the lean mass loss, adipose fibrosis, and weight regain that limit durable outcomes, particularly in older patients with established chronic obesity. ’005 is mechanistically positioned to address this gap. By inducing apoptosis of chronically activated CD206⁺ adipose macrophages and suppressing the cytokine burden generated during adipose remodeling, ’005 could synchronize the resolution of adipose inflammation with the pace of fat mass reduction. This would be expected to preserve lean mass by reducing cytokine-driven muscle catabolism, attenuate adipose fibrosis by removing the macrophage-driven profibrotic signal before it is consolidated in extracellular matrix, and reduce weight regain driven by residual low-grade inflammatory tone in reorganized adipose depots. Given that GLP-1 receptor agonists and dual GIP/GLP-1 agonists are now deployed at population scale, the potential to combine these agents with a short-course, macrophage-targeted anti-inflammatory to optimize body composition outcomes merits dedicated investigation.

Fifth, and potentially the largest near-term opportunity given existing patient populations, is the use of ’005 as an adjunct in patients who are already receiving biologic cytokine inhibitors or JAK inhibitors but have not achieved full remission. A substantial proportion of patients on anti-TNF agents, anti-IL-6 receptor antibodies, or tofacitinib-class JAK inhibitors remain in partial response: systemic inflammatory markers are reduced but not normalized, residual disease activity persists, and critically, the downstream consequences of sustained low-to-moderate inflammation continue to accumulate. These include progressive blood-brain barrier compromise, microglial activation, and neuroinflammation; ongoing vascular endothelial injury; and the HPA-to-RAAS dysregulation axis described above. Because ’005 operates through a mechanism entirely orthogonal to cytokine pathway blockade, depleting the macrophage population that generates cytokines rather than neutralizing cytokines already in solution, it can be added to existing biologic or JAK inhibitor regimens without pharmacodynamic competition or additive immunosuppression risk. A short course of ’005, administered at the cellular level where residual macrophage activation sustains the incompletely treated inflammatory state, would be expected to rapidly and broadly lower cytokine burden across multiple pathways simultaneously, including pathways not targeted by the patient’s existing therapy. Of particular clinical importance is the CNS compartment.

Chronically elevated circulating cytokines in incompletely treated systemic inflammatory disease are associated with progressive BBB dysfunction, white matter changes, and microglial priming, a state of heightened reactivity that amplifies neuroinflammatory responses to subsequent systemic insults [29,30]. Because ’005 is designed to reach CSF-adjacent compartments, a property under direct evaluation in a dedicated CSF PK study (Study 3), and targets CD206⁺ perivascular macrophages and microglia without systemic GC exposure, it may provide neuroprotection in this setting that neither systemic GCs (HPA suppression, neuropsychiatric risk) nor cytokine inhibitors (limited CNS penetration, pathway-specific coverage) can offer. Prospective trials in partially responding patients on established biologics or JAK inhibitors, with CNS imaging and cognitive endpoints alongside standard disease activity measures, would test this hypothesis directly.

The practical feasibility of adding ’005 to existing medication regimens is directly supported by the Phase II/III trial design. No exclusion criteria were applied based on comorbidities or concomitant medications, with the sole exception of prior systemic corticosteroids or immunosuppressants within 14 days of enrollment, an exclusion driven by the need to isolate the efficacy signal of each study arm, not by any concern regarding drug–drug interactions with ’005. Consequently, the 309 enrolled patients represented a heterogeneous real-world population carrying a broad range of concurrent conditions and receiving a diverse array of background medications. No drug interaction signals emerged across any concomitant drug class. This is mechanistically consistent with the pharmacokinetic profile of ’005: rapid systemic clearance with a half-life of approximately 1.9 hours, intact renal excretion of unchanged prodrug, and no detectable free dexamethasone in plasma eliminate the principal routes by which drug–drug interactions typically arise, namely, CYP450 competition, protein binding displacement, and off-target receptor engagement. Because ’005 acts intracellularly within CD206⁺ macrophages and is cleared from systemic circulation before most concomitant medications undergo their primary metabolic processing, the addition of a fixed three-day course of ’005 to a patient’s existing medication regimen should not require dose adjustment of background therapies or raise drug interaction concerns as a barrier to clinical use.

In summary, ’005 exemplifies a new class of macrophage-targeted corticosteroids that retain the unmatched speed and breadth of GC efficacy while shedding the endocrine and immunosuppressive liabilities that have limited GCs for generations. The convergence of mechanistic selectivity, prodrug integrity confirmed in two completed human PK studies, pathogen-agnostic preclinical efficacy, and replicated clinical superiority in 309 patients argues that targeted corticosteroid delivery can be reimagined from a blunt necessity into a precise, restorative intervention. Larger randomized trials across acute and chronic indications, anchored by rigorous endocrine, immune, and macrophage-biology endpoints, are now warranted.

## Materials and Methods

### Study Designs

These studies integrated mechanistic, preclinical, and clinical approaches. Mechanistic assays included receptor-binding, uptake, apoptosis, and cytokine modulation studies in human macrophages. Preclinical in vivo models spanned bacterial, viral, and parasitic infections and GLP toxicology. Clinical evaluation comprised two completed Phase I PK/safety studies and a cerebrospinal fluid PK study, and a GCP-compliant adaptive Phase II/III randomized controlled trial.

### Compound Design

’005 is a type 1a dexamethasone prodrug consisting of a mannose moiety conjugated via an oxime-ether, pH-sensitive linker to dexamethasone (MW 627 Da). Designed for selective CD206-mediated uptake with low micromolar binding affinity, recycling compatibility, and intracellular release triggered by endosomal acidification. Lyophilized formulation reconstituted in 5 mL 0.9% NaCl (4 mg/mL) for IV bolus administration. Administered under yellow monochromatic light with patient supine.

### Binding and Uptake Assays

Recombinant human CD206 (His-tagged; Acro Biosystems) was used to quantify binding affinity by microscale thermophoresis (MST) and surface plasmon resonance (SPR). FITC-labeled ’005 was incubated with primary human monocyte-derived macrophages polarized to M0 or M2 phenotypes. Uptake was assessed by flow cytometry with trypan blue quenching to distinguish surface-bound from internalized compound.

### Apoptosis Assays

Human M2 macrophages were cultured under serum-free conditions with 1.2 mM Ca²⁺ and 0.5 mM Mg²⁺, then treated with ’005 (25–100 µg/mL) or vehicle for 48 h. Cell viability was assessed by Annexin V/propidium iodide staining and flow cytometry.

### Cytokine Assays in SARS-CoV-2–Infected Macrophages

Human monocyte-derived macrophages were transfected with ACE2 mRNA and infected with SARS-CoV-2 (MOI = 5). Cells were treated with ’005 (10 µg/mL), dexamethasone (10 nM), or vehicle.

Supernatants were collected at 24 h and analyzed for IL-6, TNF, and CXCL10 by ELISA.

### In Vitro and In Vivo Leishmania Assays

In vitro: THP-1–derived macrophages infected with GFP-tagged L. donovani promastigotes (5×10⁶/well); after 36 h, treated with ’005 (0.1–10 µg/mL). Parasite load and viability quantified 12–14 h later by fluorescence microscopy. In vivo visceral leishmaniasis: BALB/c mice infected with L. donovani (5×10⁵) treated IV with 100 µg ’005; parasite burden quantified at Day 15. Cutaneous leishmaniasis: BALB/c mice infected with L. major received five subcutaneous doses of ’005; lesion size monitored through Day 60.

### 28-Day GLP Rat Toxicology

Male Wistar rats (n = 6/group) received daily IV ’005 (10, 20, or 30 mg/kg), dexamethasone (3–10 mg/kg), or saline for 28 days. Histopathology of liver, lung, and bone was performed by blinded pathologists. Serum chemistry and hematology panels were measured weekly. NOAEL = 30 mg/kg.

### Human Pharmacokinetics

Study 1 — First-in-human dose-escalation and repeated-dose study (PIF-GC-CP-101; NCT05619770; Rambam Health Care Campus, Haifa, Israel): Phase I, single-center, open-label study in healthy adult volunteers aged 18–50 years (BMI ≤ 30), of whom 15 received ’005. A single-dose escalation arm administered 10, 20, and 30 mg IV bolus (one dose per cohort); a repeated-dose arm (n = 3) received 30 mg IV once daily for 3 consecutive days. Free plasma dexamethasone was quantified by LC-MS/MS.

Morning cortisol and ACTH were measured at screening and 24 h after the final dose to assess HPA-axis function. Safety endpoints included adverse events, hematology, biochemistry, urinalysis, vital signs, ECG, and blood glucose.

Study 2 — Dedicated single-/multiple-dose PK and safety study (ICS/LAX/2022-004; ICBio Clinical Research, Bangalore, India): single-arm, non-randomized, open-label study in 6 healthy adult male subjects. Subjects received 30 mg ’005 IV bolus as a single dose and then once daily for 3 consecutive days under fasting conditions while recumbent and under yellow monochromatic light. Plasma for intact ’005 and free dexamethasone was collected within 30 min pre-dose and at 0.5, 0.75, 1, 1.5, 2, 3, 6, 9, 12, and 24 h post-dose on Day 1 and Day 3. Urine was collected after Day 1 dosing over 0–4, 4–8, 8–12, 12–18, and 18–24 h intervals for unchanged ’005. Analytes were quantified by validated LC-MS/MS. PK parameters included Cmax, Tmax, t1/2, AUC0-t, AUC0-inf, Kel, and urinary Rmax/AE24; descriptive statistics were generated in SAS. Safety monitoring included local and systemic AEs, hematology, biochemistry, urinalysis, physical examination, vital signs, ECG, random blood glucose, and injection-site tolerability.

Study 3 — Cerebrospinal fluid PK study (ICS/LAX/2023-002; ICBio Clinical Research, Bangalore, India): single-arm, open-label, single-dose study in 6 healthy adult subjects designed to characterize CSF and plasma PK, safety, and tolerability of a 30 mg IV bolus of ’005. CSF was obtained by serial lumbar sampling (approximately 1.5 mL per sample) within 45 min pre-dose and at 0.75, 1.5, 2.5, 3.5, 6.5, 9.5, 12.5, and 24.5 h post-dose; plasma was sampled within 30 min pre-dose and at 0.5, 1, 2, 3, 6, 9, 12, and 24 h post-dose. ’005 and free dexamethasone were quantified by an LC-MS/MS method validated in human CSF over 0.02–4 ng/mL. The clinical study report was not yet finalized at the time of writing.

### Phase II/III Clinical Trial Design (NCT05656521; Protocol ICS/LAX/2022-005)

Prospective, randomized, open-label, active-controlled adaptive Phase II/III trial. Phase II: n = 62 (31+31), 4 sites, January–April 2023. DSMB interim review approved Phase III continuation. Phase III: n = 247 (124+123), 6 sites (same 4 + 5 new), January 2024–July 2025. Total enrolled: 309 (155 ’005 + SOC; 154 Dexa + SOC). Eligible: adults with moderate COVID-19, WHO score 4–5, supplemental oxygen without mechanical ventilation. No comorbidity restrictions. Randomization 1:1; no blinding (open-label active-control design). Protocol allowed investigator-initiated supplementary medications at any time. Primary endpoint: time to ≥2-point WHO ordinal scale improvement. Non-inferiority margin: HR 0.70. Pre-specified secondary endpoints: viral clearance by RT-PCR, hospital discharge, SpO₂, body temperature, CRP/IL-6/D-dimer, all-cause mortality. Follow-up to Day 28 (HRCT lung assessment). All trials approved by CDSCO and by independent Ethics Committees at each participating site; all patients provided written informed consent.

## Statistical Analysis

Data are presented as mean ± SD or SEM. In vitro and animal comparisons: two-tailed unpaired t-tests or two-way ANOVA with post hoc correction; categorical variables by Fisher’s exact test. Clinical trial: Kaplan–Meier analysis for time-to-event endpoints; hazard ratios and 95% CIs by Cox proportional hazards model. Categorical outcomes by Fisher’s exact test. Continuous longitudinal outcomes by ANCOVA or MMRM. Two-sided p < 0.05 considered statistically significant. All clinical analyses conducted using SAS 9.4.

## Data Availability

All data produced in the present study are available upon reasonable request to the authors

## Acknowledgments

We thank the healthy volunteers and patients who participated in the Phase I and Phase II/III clinical studies, and the investigators and clinical staff at all participating sites for their dedication and care. We acknowledge Dr. Avivit Peer (Rambam Medical Center) for leadership of the Phase I studies, and the clinical investigators at the six Phase II/III sites (Pune, Mumbai, Nellore, Bengaluru, Gandhinagar) for their conduct of the trial. We thank Dr. Ramachandran Murali (Cedars-Sinai Medical Center) for CD206 binding studies, Dr. Shulamit Michaeli (Bar-Ilan University) for leishmaniasis models, and Dr. Larissa Labzin (University of Queensland) for SARS-CoV-2 macrophage assays. We also thank Vivox Ltd. (Nesher, Israel), Laxai Life Sciences (Hyderabad, India), Insignia Clinical Services (Delhi, India), and the La Jolla Institute for Immunology (San Diego, USA) for preclinical in vivo studies and clinical operations support.

## Author contributions

Conceptualization: MMG. Methodology: AMG, OW. Visualization: JIG. Funding acquisition: MMG, AMG, JIG. Writing – original draft: MMG. Writing – review and editing: JIG, AMG, OW.

## Data and materials availability

All data supporting the findings of this study are available within the main text or the Supplementary Materials. The Phase II/III CSR (Protocol ICS/LAX/2022-005) and the Phase I study reports (PIF-GC-CP-101; ICS/LAX/2022-004; and the CSF study ICS/LAX/2023-002, report pending) are available from the corresponding author on reasonable request. ClinicalTrials.gov: NCT05656521 (Phase II/III), NCT05619770 (first-in-human Phase I).

## Supplemental Materials and Methods

### Systemic inflammation

C57BL/6 mice received intraperitoneal LPS (5 mg/kg). Thirty minutes later, animals were treated IV with ‘005 (1 mg), dexamethasone (0.45 mg/kg), or PBS. Clinical scores were assessed at 24 h by blinded observers.

### Bacterial sepsis

C57BL/6 mice were infected intraperitoneally with *Streptococcus pneumoniae* (10³ CFU). Animals received IV ‘005 (3 mg) or vehicle at 2 h and 24 h. CFU counts in peritoneal fluid and blood were measured at 48 h.

### Dengue virus challenge

AG129 mice were pretreated with anti-DENV mAb 2H2 and infected with DENV2 S221. Mice received IV ‘005 (0.4 mg) or PBS at 24, 48, and 72 h. Survival was monitored for 10 days.

## Supplemental Figures

**Suppl. Fig. 1.**
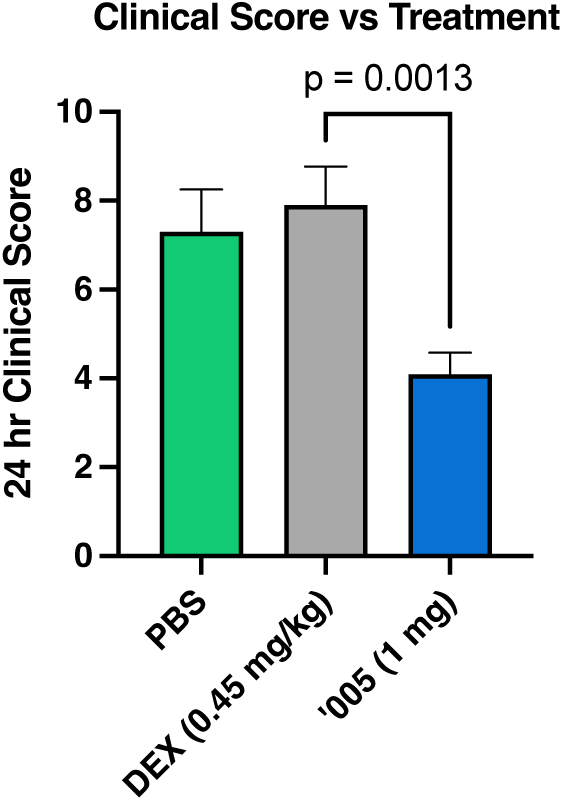
Clinical score in systemic LPS challenge model. Clinical score at 24 h following systemic LPS challenge in C57BL/6 mice treated with vehicle (PBS), dexamethasone (0.45 mg/kg), or ‘005 (1 mg). Clinical scores reflect combined assessment of physical appearance, response to stimulus, eye condition, and respiration (higher scores indicate worse condition). Bars represent mean ± SEM. *P* = 0.0013 for 101-PGC-005 vs dexamethasone (one-way ANOVA).

**Suppl. Fig. 2.**
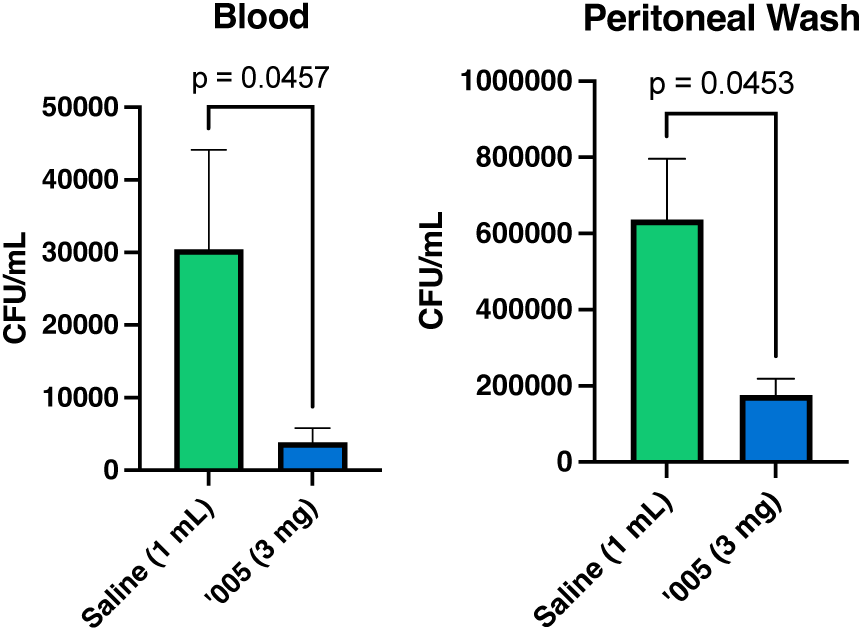
Bacterial sepsis model. (a) Bacterial burden in peritoneal wash at 48 h post-infection following intraperitoneal challenge with 10³ CFU of S. pneumoniae strain INV104B and treatment with 101-PGC-005 (3 mg in saline, IV, at 2 h and 24 h post-infection; n = 10) or saline control (n = 10). Bars show mean ± SEM; p = 0.0453 by unpaired t-test. (b) Bacterial burden in blood from the same animals, p = 0.0457 by unpaired t-test.

**Suppl. Fig. 3.**
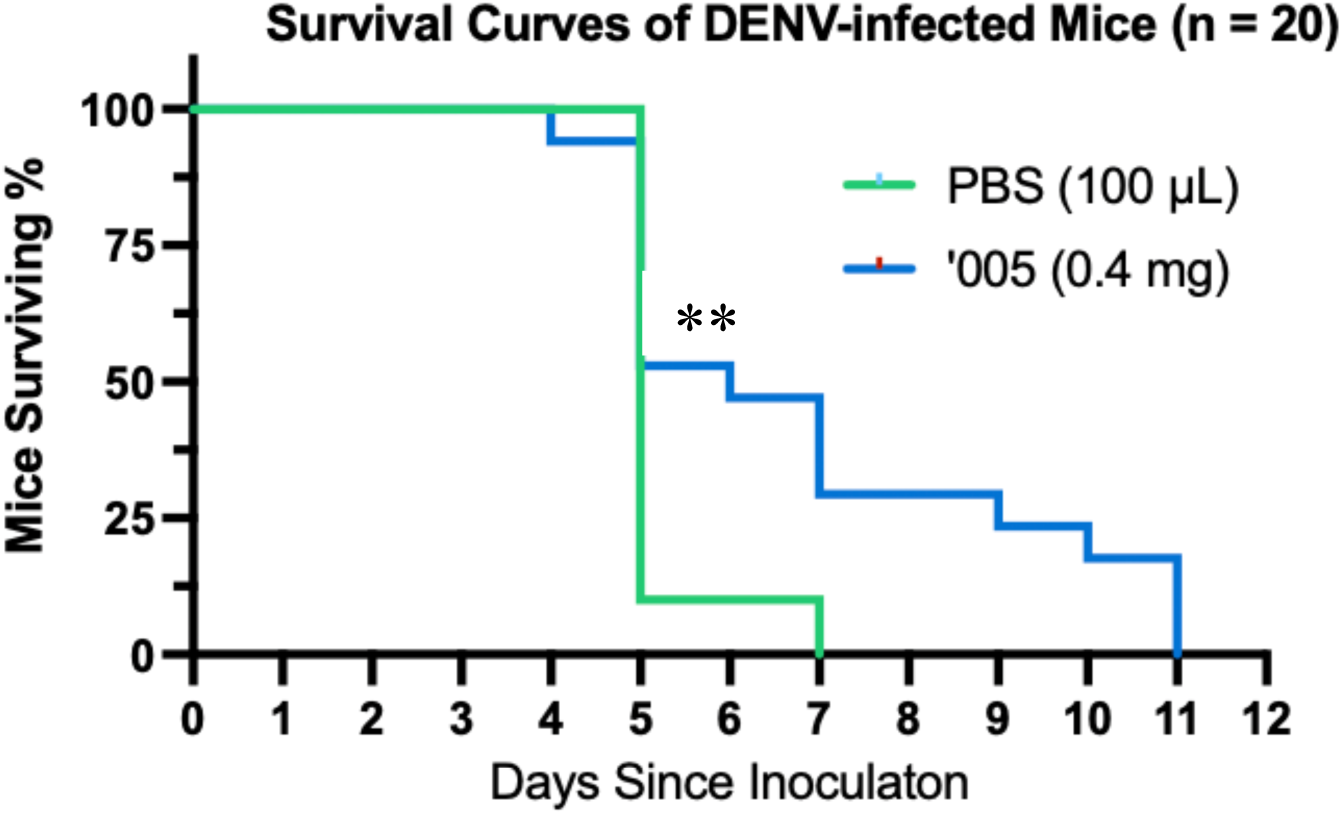
Dengue survival model. Kaplan–Meier survival curves for AG129 mice (n = 20 per group) infected with DENV2 strain S221 via antibody-dependent enhancement (ADE) and treated with either 101-PGC-005 (0.4 mg in 100 μL PBS, retro-orbital injection) or PBS control. Treatment was administered once daily at 24-, 48-, and 72-hours post-infection. Mice were monitored for 10 days and euthanized upon ≥20% weight loss or a clinical score of 5. Survival was significantly prolonged in 101-PGC-005–treated mice compared with PBS-treated controls (log-rank test, p < 0.05).

